# Too early for LATE? Age at onset is associated with the clinical and neuroimaging expression of limbic-predominant age-related TDP-43 pathology in Alzheimer’s disease

**DOI:** 10.64898/2026.10.15.26365384

**Authors:** Nuole Zhu, Renaud La Joie, Isabel Elaine Allen, Marlene Lin, D. Luke Fischer, David N. Soleimani-Meigooni, Howard J. Rosen, Maria Luisa Gorno-Tempini, Zachary Miller, David C. Perry, Joel Kramer, Katherine L. Possin, Yann Cobigo, Lyzzette Melgoza, Argentina Lario Lago, Jennifer Yokoyama, Claire Yballa, Ganna Blazhenets, Bruce L. Miller, Lea T. Grinberg, William W. Seeley, Salvatore Spina, Gil D. Rabinovici

## Abstract

Limbic-predominant age-related TDP-43 encephalopathy neuropathologic change (LATE-NC) frequently co-occurs with Alzheimer’s disease (AD) pathology and has been studied mainly in older populations. We examined its frequency and expression in 196 individuals with autopsy-confirmed intermediate-to-high AD neuropathologic change, enriched for younger-onset cases, integrating neuropathology with antemortem clinical, cognitive and neuroimaging data. LATE-NC occurred in 31% overall and 19% of individuals with symptom onset ≤65 years. Its frequency increased with older age at symptom onset, and its presence was associated with greater episodic memory impairment and smaller amygdala and hippocampal volumes. Among individuals with LATE-NC, older age at symptom onset was associated with memory-first presentation and greater medial temporal involvement, including higher inferior-to-medial temporal metabolism ratios on FDG PET. Despite its lower frequency, LATE-NC co-pathology should still be considered in early-onset AD, where selected clinical features and neuroimaging markers described in older populations may be less applicable.

## Introduction

Decades of post-mortem investigations of individuals with cognitive decline have established that Alzheimer’s disease neuropathological change (ADNC) rarely occurs in isolation: mixed neuropathological findings are the norm in the aging brain.^1,2^ Limbic-predominant age-related TDP-43 encephalopathy neuropathologic change (LATE-NC) has emerged as one of the most frequent and clinically relevant modifiers of the Alzheimer’s disease (AD) phenotype. Community-based autopsy studies have shown that LATE-NC is present in more than 20% of individuals over 80 years of age, with prevalence increasing markedly with advancing age and in the presence of ADNC.^3–5^ The coexistence of LATE-NC and ADNC has been associated with worse cognitive performance across multiple domains, particularly episodic memory, and a more aggressive clinical course than either pathology alone.^3,5–9^ As concomitant neuropathological entities are increasingly recognized as major contributors to the clinical heterogeneity of AD,^10^ understanding the impact of LATE-NC and identifying affected individuals during life have become a priority, particularly as co-pathology not targeted by AD disease-modifying therapies may influence therapeutic response, with implications for weighing treatment risks against benefits.

Despite its growing clinical relevance, no validated molecular biomarker is currently available to detect TDP-43 proteinopathy during life. Consequently, considerable efforts have focused on recognizing LATE-NC using combinations of clinical features and neuroimaging markers. These studies have shown that LATE-NC leaves a relatively characteristic clinical and neuroimaging signature, even in the presence of concomitant AD neuropathology. Compared with AD alone, individuals with concomitant LATE-NC consistently exhibit disproportionately greater episodic memory impairment together with preferential involvement of limbic structures, particularly within the medial temporal lobe, on structural MRI and [^18^F]fluorodeoxyglucose PET (FDG PET).^3,6–9,11–14^ This pattern of disproportionate medial temporal involvement relative to neocortical neurodegeneration has motivated the development of regional neuroimaging ratios, including entorhinal-to-parahippocampal (ERC/PHC) volume ratios on MRI and inferior-to-medial temporal (IMT) metabolism ratios on FDG PET, to improve the *in vivo* identification of LATE-NC.^12,13,15,16^ More recently, these observations have culminated in consensus clinical criteria for LATE, providing an initial framework to identify individuals likely to harbor LATE-NC during life.^17^ In individuals with biomarker-confirmed AD, these criteria support a diagnosis of possible concomitant LATE when medial temporal impairment is disproportionate to clinical disease severity or tau burden, or when FDG PET shows an elevated IMT metabolism ratio.^17^ However, nearly all evidence supporting these clinical and neuroimaging frameworks has been derived from groups of individuals with late symptom onset, typically those aged 75 years and older.

Although LATE-NC has been regarded as an age-related pathology, accumulating neuropathological evidence indicates that it is not restricted to late-onset disease. LATE-NC has been reported as a co-pathology in autosomal dominant Alzheimer’s disease (ADAD), Down syndrome-associated Alzheimer’s disease (DSAD), and autopsy cohorts enriched for sporadic early-onset Alzheimer’s disease (EOAD). Reported frequency in these younger populations ranges between 10% and 25%, although many estimates are based on autopsy samples of only a few dozen individuals.^18–22^ Existing evidence in these populations is also largely limited to post-mortem neuropathological characterization, leaving its *in vivo* clinical and neuroimaging expression and its influence on disease progression poorly understood.^18–21,23^ Consequently, it is unclear whether the limbic-predominant phenotype and the clinical-neuroimaging frameworks established in older populations remain applicable to younger individuals.

In this study, we examined the frequency of LATE-NC and its clinical and neuroimaging expression across the AD age-at-onset continuum using a large single-center autopsy-confirmed cohort enriched for EOAD. We included clinically impaired individuals from the University of California, San Francisco (UCSF) Neurodegenerative Disease Brain Bank (NDBB) with intermediate-to-high ADNC and excluded cases with frontotemporal lobar degeneration with TDP-43 (FTLD-TDP) or tau (FTLD-tau) pathology. By relating post-mortem LATE-NC status to longitudinal clinical, neuropsychological, neuroimaging and genetic data acquired during life, we asked how often LATE-NC occurs across this continuum, how it relates to these measures, and whether its expression differs by age at symptom onset. We hypothesized that LATE-NC would be present throughout the age-at-onset continuum, with increasing frequency at older ages of symptom onset, and that it would retain its limbic-predominant signature even in younger-onset cases, with greater episodic memory impairment and medial temporal involvement than in cases without LATE-NC.

## Results

### Clinical characteristics and LATE-NC frequency across the age at symptom onset continuum

The study cohort comprised 196 individuals, of whom 31% (61) had LATE-NC (stages 1-3) (Figure 1 and Table 1). Individuals with LATE-NC had older age at symptom onset (67.9 ± 9.4 vs. 60.2 ± 10.1 years, *P* < 0.001) and death (80.9 ± 8.2 vs. 70.9 ± 10.3 years, *P* < 0.001). Disease duration from symptom onset to death was also longer in the LATE-NC group (12.8 ± 4.7 vs. 10.3 ± 3.8 years, *P* < 0.001). Sex distribution was similar between groups. Participants with LATE-NC were slightly more highly educated (17.8 ± 3.1 vs. 16.5 ± 3.5 years, *P* = 0.007) and had higher baseline Mini-Mental State Examination (MMSE) scores (23.8 ± 5.6 vs. 21.2 ± 6.1, *P* = 0.008), whereas baseline Clinical Dementia Rating-Sum of Boxes (CDR-SB) scores did not differ between groups. Demographic, clinical, and neuropathological differences remained largely consistent when analyses were restricted to early-onset cases, defined as symptom onset at age 65 years or younger (Supplementary Table 1).

**Figure 1.**
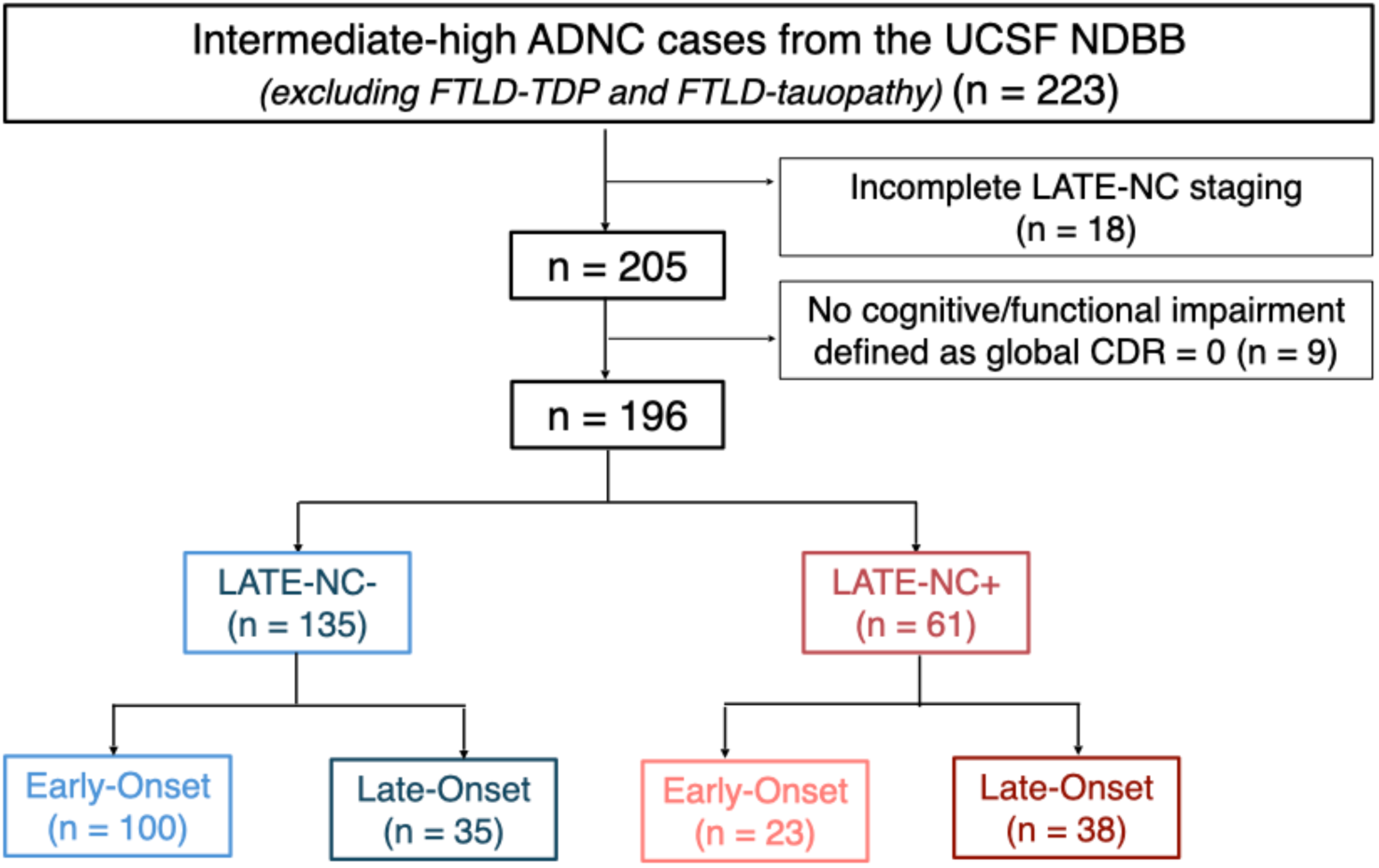
Flow chart of patient selection. ADNC = Alzheimer’s disease neuropathologic change; FTLD, frontotemporal lobar degeneration; TDP, transactive response DNA-binding protein 43 kDa; LATE-NC, limbic-predominant age-related TDP-43 encephalopathy neuropathologic change; CDR, Clinical Dementia Rating.

**Table 1.** Clinical and neuropathologic characteristics according to LATE-NC status.

Tables, Figures and Legends
|  |  |  |  |
| --- | --- | --- | --- |
|  | LATE-NC-<br>(n = 135) | LATE-NC+<br>(n = 61) | P-value |
| **Demographic and clinical characteristics** |  |  |  |
| Age at symptom onset, years | 60.2 ± 10.1 | 67.9 ± 9.4 | <0.001 |
| Age at death, years | 70.9 ± 10.3 | 80.9 ± 8.2 | <0.001 |
| Duration from symptom onset to death, years | 10.3 ± 3.8 | 12.8 ± 4.7 | <0.001 |
| Female sex, n (%) | 64 (47%) | 31 (51%) | 0.76 |
| Education, years | 16.5 ± 3.5 | 17.8 ± 3.1 | 0.007 |
| Clinical phenotype, n (%) |  |  | 0.022 |
| Amnesic | 73 (59%) | 45 (80%) |  |
| Non-amnesic* | 16 (13%) | 6 (11%) |  |
| Primary progressive aphasia | 17 (14%) | 2 (3.6%) |  |
| Posterior cortical atrophy | 18 (15%) | 3 (5.4%) |  |
| Baseline MMSE | 21.2 ± 6.1 | 23.8 ± 5.6 | 0.008 |
| Baseline CDR Sum of Boxes | 4.6 ± 2.5 | 4.3 ± 3.2 | 0.60 |
| **Genetic characteristics** |  |  |  |
| APOE ε4 carrier, n (%) | 60/123 (49%) | 31/52 (60%) | 0.25 |
| TMEM106B rs1990622 A/A, n (%) | 36/119 (30%) | 24/52 (46%) | 0.056 |
| Both APOE ε4 carrier and TMEM106B rs1990622 A/A, n (%) | 20/119 (17%) | 16/52 (31%) | 0.044 |
| **Neuropathologic features** |  |  |  |
| AD neuropathologic change (ADNC)**, n (%) |  |  | 1.00 |
| Intermediate | 10 (7%) | 5 (8%) |  |
| High | 125 (93%) | 55 (92%) |  |
| Lewy body disease, n (%) | 75 (56%) | 33 (54%) | 0.97 |
| Hippocampal sclerosis, n (%) | 4 (3%) | 16 (28%) | <0.001 |
| Cerebral amyloid angiopathy, n (%) | 107 (79%) | 53 (88%) | 0.16 |
| Vascular pathology, n (%) | 46 (34%) | 25 (41%) | 0.41 |

Data are presented as mean ± standard deviation for continuous variables and n (%) for categorical variables. Between-group comparisons were performed using Student's t-test for continuous variables and $\chi^2$ test for categorical variables. Genetic variables were compared using Fisher's exact test. Percentages were calculated based on available data; denominators may vary because of missing data. \*Non-amnesic phenotype included dysexecutive and other atypical presentations not classified as primary progressive aphasia or posterior cortical atrophy. \*\* One LATE-NC+ case had ADNC severity indeterminate between intermediate and high.
AD, Alzheimer's disease; APOE, apolipoprotein; LATE-NC, limbic-predominant age-related TDP-43 encephalopathy neuropathologic change; TMEM106B, transmembrane protein 106B.

Genetic screening identified two *APP* mutation carriers and one individual carrying a possibly pathogenic *PSEN1* variant, all without LATE-NC. No pathogenic variants were identified in *PSEN2*, *MAPT*, *GRN*, *TARDBP* or *FUS*, and no *C9orf72* expansions were detected. *APOE* ε4 carrier status and the *TMEM106B* rs1990622 A/A genotype were both more frequent among individuals with LATE-NC, although neither difference reached statistical significance (*APOE* ε4: 60% vs. 49%, P = 0.25; *TMEM106B* A/A: 46% vs. 30%, P = 0.056). By contrast, co-occurrence of *APOE* ε4 and the *TMEM106B* A/A was significantly higher in the LATE-NC group (31% vs. 17%, P = 0.044) (Table 1). Similar patterns were observed when analyses were restricted to individuals with early-onset disease (Supplementary Table 2).

LATE-NC was present in 19% (23/123) of individuals with symptom onset at or before age 65, but its frequency increased markedly thereafter, reaching 52% (38/73) (Figure 1 and 2A). When stratified by age at death, only one individual (2%) who died at or before age 65 had LATE-NC. The frequency increased to 23% (13/56) in individuals who died between 66 and 75 years and to approximately 54% (47/87) in those who died at age 76 or older (Figure 2B and Supplementary Table 3). Among individuals with LATE-NC, stage 2 pathology (hippocampal/entorhinal involvement) predominated (79%, 48/61), whereas only one individual (age of onset = 66-75 years, age at death ≥ 86 years) reached stage 3 (middle frontal gyrus involvement). Higher LATE-NC stage (stage 2-3 vs. stage 1) was not associated with age at symptom onset, age at death, or disease duration (ORs = 0.96 to 1.14, all 95% CIs included 1). ADNC severity and the frequency of major co-pathologies, including Lewy body disease, cerebral amyloid angiopathy, and vascular pathology, were comparable between LATE-NC groups, whereas hippocampal sclerosis was substantially more frequent in individuals with LATE-NC (28%, 16/57 vs. 3%, 4/135; P < 0.001) (Table 1).

**Figure 2.**
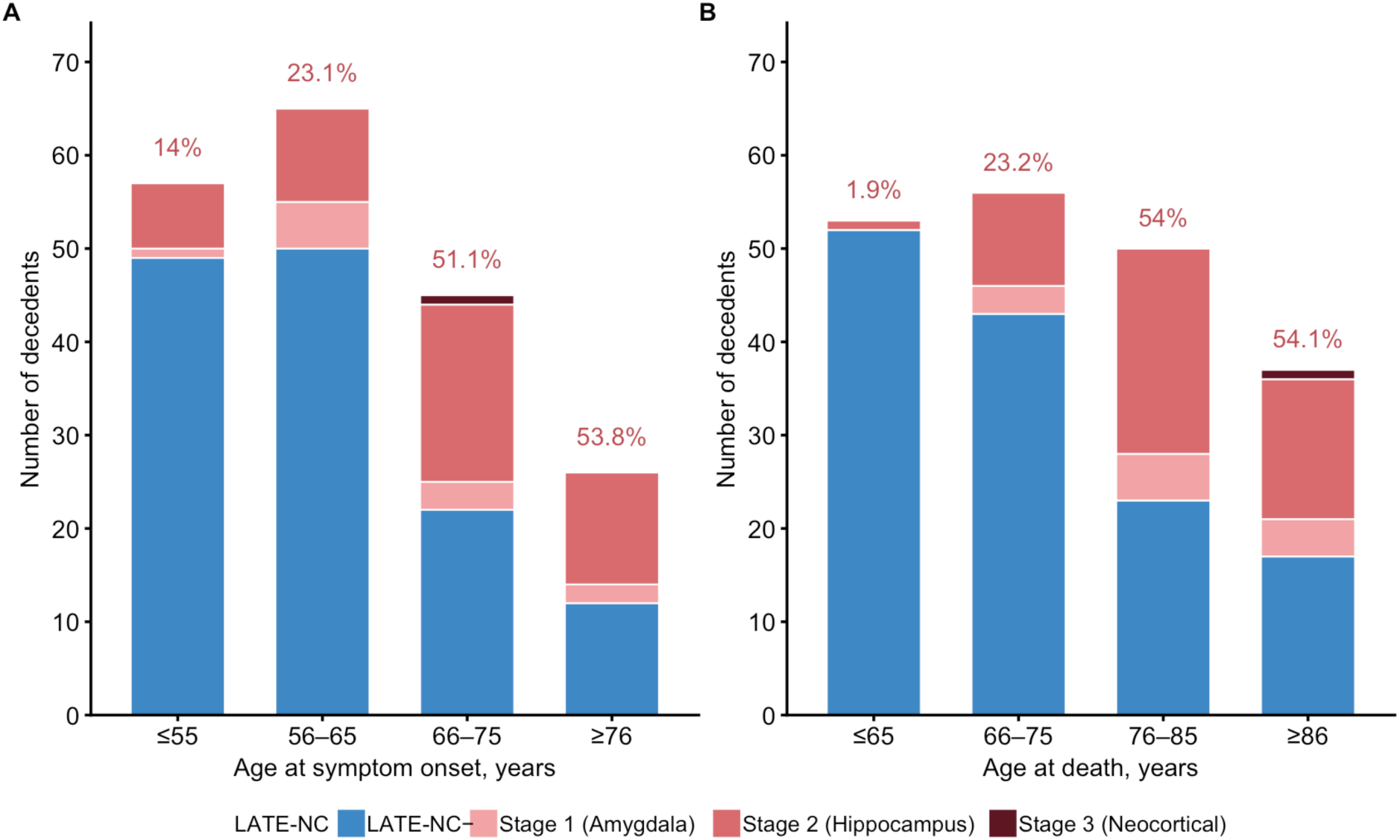
LATE-NC stage distribution according to age at symptom onset and age at death. Distribution of LATE-NC stages stratified by (A) age at symptom onset and (B) age at death. Stacked bars represent the number of decedents without LATE-NC (LATE-NC-) and with LATE-NC stages 1-3 within each group. Percentages above the bars indicate the proportion of decedents with LATE-NC (stages 1-3 combined) within each age group. LATE-NC, limbic-predominant age-related TDP-43 encephalopathy neuropathologic change.

### LATE-NC is associated with a more amnestic phenotype, but the likelihood of a memory-first presentation depends on age at symptom onset

At first evaluation, 80% (45/56) of individuals with LATE-NC presented with an amnestic phenotype compared with 59% (73/124) of those without LATE-NC (OR = 2.86, 95% CI 1.41 to 6.78) (Figure 3A). Individuals with LATE-NC were also more likely to report memory as the first presenting symptom (OR = 3.46, 95% CI 1.78 to 7.44). The association between age at symptom onset and memory as first symptom differed by LATE-NC status (interaction OR = 2.44 per 10-year increase, 95% CI 1.04 to 8.88). Among individuals without LATE-NC, age was not associated with memory-first presentation (OR = 0.91 per 10-year increase, 95% CI 0.50 to 1.45), whereas among those with LATE-NC, older age was associated with an increased likelihood of memory as the presenting symptom (OR = 2.21 per 10-year increase, 95% CI 1.08 to 7.18) (Figure 3B).

**Figure 3.**
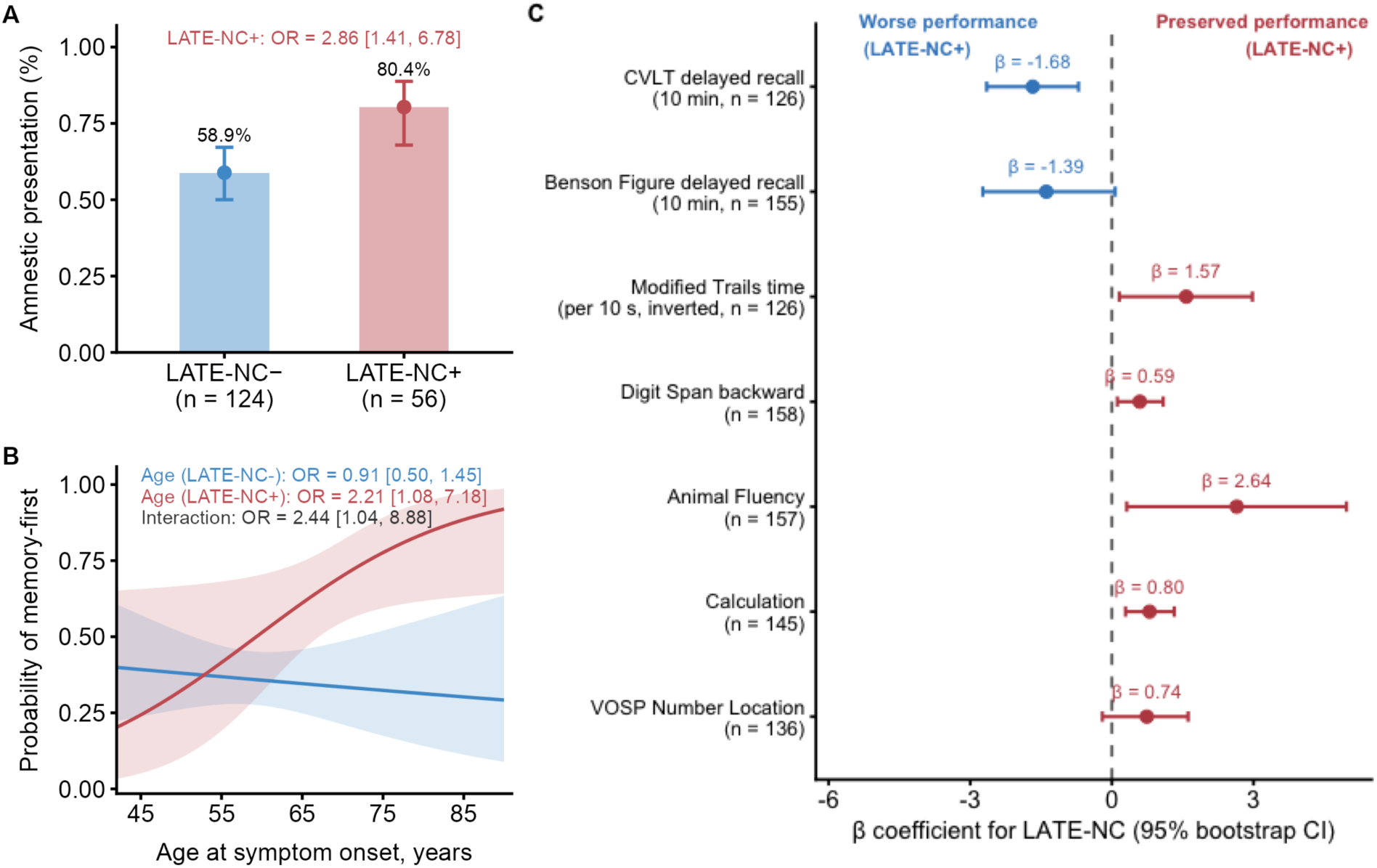
Clinical presentation and cognitive performance according to LATE-NC status. (A) Observed proportion of individuals with an amnestic presentation at first evaluation according to LATE-NC status. Bars represent observed proportions, points and error bars represent model-estimated probabilities and their 95% CIs. (B) Model-estimated probability of memory being the first reported symptom across age at symptom onset, stratified by LATE-NC status. Shaded areas represent 95% CIs. (C) Associations between LATE-NC and cognitive performance at first evaluation. Points represent β coefficients for LATE-NC and error bars represent 95% CIs. Modified Trails time was inverted, so higher values indicate better performance. Models were adjusted for age at symptom onset, with sex and education retained when they improved model fit, 95% CIs were estimated using 5,000 bootstrap resamples. The Boston Naming Test was also evaluated, but LATE-NC was not retained in the final AIC-selected model and therefore is not shown. CI, confidence interval; CVLT, California Verbal Learning Test; LATE-NC, limbic-predominant age-related TDP-43 encephalopathy neuropathologic change; VOSP, Visual Object and Space Perception.

At the first available neuropsychological assessment, LATE-NC was associated with a relatively selective episodic memory impairment, with worse delayed verbal recall on the California Verbal Learning Test-Short Form (CVLT) at 10 minutes (β = −1.68, 95% CI −2.65 to −0.71), while delayed visual recall on the Benson Figure at 10 minutes showed a similar but non-significant trend (β = −1.39, 95% CI −2.69 to 0.06). In contrast, performance across non-memory cognitive domains was largely preserved, with executive function, attention, animal fluency and calculation scores generally showing better performance among individuals with LATE-NC (Figure 3C). No association with LATE-NC was observed for the Boston Naming Test.

After excluding stage 1 (amygdala-only involvement), sensitivity analyses comparing LATE-NC stages 2-3 with stage 0 yielded consistent findings across clinical phenotypes, presenting symptoms, and cognitive performance. Individuals with LATE-NC stage 2-3 remained more likely to present with an amnestic clinical phenotype (OR = 2.38, 95% CI 1.10 to 6.02) and to report memory as the first symptom (OR = 2.85, 95% CI 1.35 to 6.35) (Supplementary Figure 1A). For memory-first presentation, the effect estimate for the interaction between LATE-NC status and age at symptom onset was comparable to that in the primary analysis (interaction OR = 2.32 per 10-year increase, 95% CI 0.93 to 15.47) (Supplementary Figure 1B). Likewise, the baseline neuropsychological profile remained unchanged, showing selective impairment in CVLT delayed recall at 10 minutes (β = −1.76, 95% CI −2.73 to −0.64), together with a similar non-significant trend for Benson Figure delayed recall at 10 minutes (β = −1.22, 95% CI −2.75 to 0.37) (Supplementary Figure 1C).

When analyses were restricted to individuals with early-onset disease, LATE-NC remained associated with an amnestic clinical phenotype (OR = 3.01, 95% CI 1.08 to 16.56) and selective impairment in delayed verbal recall (CVLT delayed recall at 10 minutes: β = −1.93, 95% CI −3.17 to −0.74) (Supplementary Figure 2A-B).

### LATE-NC is associated with lower gray matter volumes in limbic regions, whereas previously proposed ERC/PHC ratio shows limited discriminative power in early-onset cases

In cross-sectional analyses of the 116 participants with structural MRI, LATE-NC was associated with a limbic-predominant pattern of lower gray matter volumes, including smaller amygdala (β = −53.9 mm^3^, 95% CI −100.1 to −7.7; area under the curve [AUC] = 0.70, 95% CI 0.58 to 0.81) and hippocampal volumes (β = −151.3 mm^3^, 95% CI −271.1 to −32.3; AUC = 0.73, 95% CI 0.61 to 0.84), after adjusting for age at symptom onset and total intracranial volume (TIV) (Figure 4A-B). Regional cortical analyses also showed lower volumes in the bilateral entorhinal (left: β = −48.8 mm^3^, 95% CI −111.7 to 9.9; right: β = −60.4 mm^3^, 95% CI −123.3 to 4.6) and medial orbitofrontal cortices (left: β = −54.1 mm^3^, 95% CI −152.4 to 44.5; right: β = - 56.9 mm^3^, 95% CI −153.5 to 45.3), although their confidence intervals included zero (Figure 4E).

**Figure 4.**
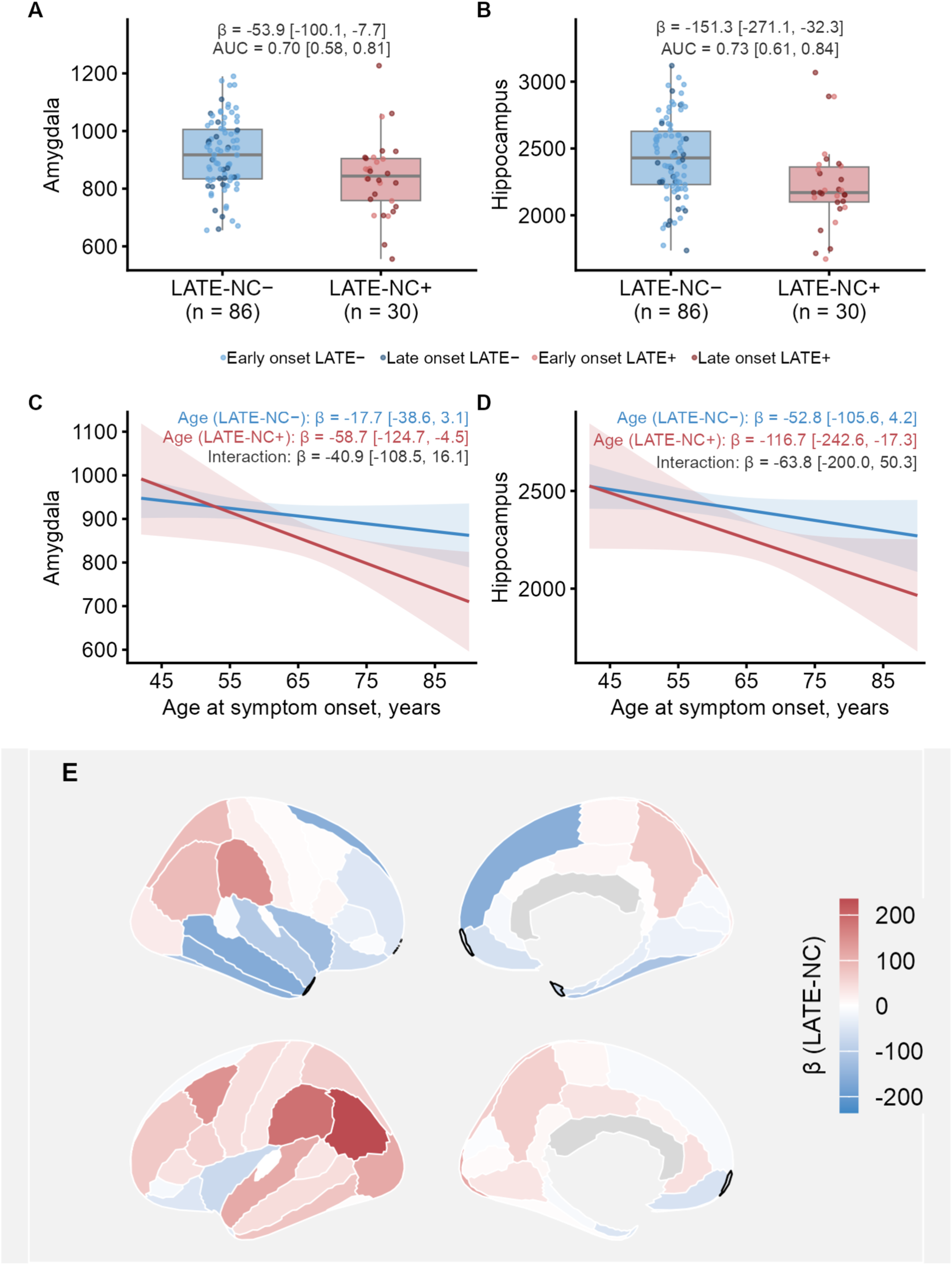
Regional gray matter volumes according to LATE-NC status and age at symptom onset. (A) Amygdala and (B) hippocampal gray matter volumes by LATE-NC status. Boxplots show the median and interquartile range, with individual observations stratified by early- and late-onset cases. β coefficients (95% CI) correspond to the association between LATE-NC and regional volumes; AUC values reflect the discriminative performance of amygdala and hippocampal volumes for LATE-NC status. (C) Amygdala and (D) hippocampal model-estimated volumes across age at symptom onset, by LATE-NC status. Lines represent model estimates and shaded areas represent 95% CIs. (E) Cortical gray matter volume differences associated with LATE-NC (LATE-NC+ vs. LATE-NC-). Upper and lower views correspond to the right and left hemispheres, respectively. Dark outlines mark regions where the 95% CIs did not include zero. Models were adjusted for age at symptom onset and total intracranial volume, with 95% CIs estimated using 5,000 bootstrap resamples. AUC, area under the curve; CI, confidence interval; LATE-NC, limbic-predominant age-related TDP-43 encephalopathy neuropathologic change.

In interaction analyses, every 10-year increase in age at symptom onset was associated with lower amygdala (β = −58.7 mm^3^, 95% CI −124.7 to −4.5) and hippocampal (β = −116.7 mm^3^, 95% CI −242.6 to −17.3) volumes within the LATE-NC group (Figure 4C-D). By comparison, the previously proposed ERC/PHC ratio did not differ according to LATE-NC status (β = - 0.04, 95% CI −0.08 to 0.01; AUC = 0.60, 95% CI 0.46 to 0.72) and the association with age at symptom onset among individuals with LATE-NC did not reach statistical significance (β = - 0.06 per 10-year increase, 95% CI −0.15 to 0.02) (Supplementary Figure 3A, D).

After exclusion of LATE-NC stage 1, associations with smaller amygdala (β = −78.8 mm^3^, 95% CI −135.9 to −22.7; AUC = 0.75, 95% CI 0.62 to 0.87) and hippocampal volumes (β = −176.8 mm^3^, 95% CI −326.5 to −30.3; AUC = 0.73, 95% CI 0.60 to 0.85) remained evident (Supplementary Figure 4A-B). Regional cortical analyses continued to show a limbic-predominant pattern of lower gray matter volume, including the bilateral entorhinal cortices (left: β = −75.2 mm^3^, 95% CI −157.7 to 0.29; right: β = −86.5 mm^3^, 95% CI −161.3 to −10.9), with similar trends in medial orbitofrontal cortices (left: β = −81.6 mm^3^, 95% CI −199.3 to 33.8; right: β = −75.3 mm^3^, 95% CI −189.1 to 38.6) (Supplementary Figure 4E). In interaction analyses, older age at symptom onset remained associated with lower amygdala and hippocampal volumes within the LATE-NC stage 2-3 group, although confidence intervals included zero (Supplementary Figure 4C-D). The ERC/PHC ratio was lower in individuals with LATE-NC (β = −0.06, 95% CI −0.12 to −0.01; AUC = 0.64, 95% CI 0.45 to 0.77), while its association with age at symptom onset was not significant (β = −0.04, 95% CI −0.18 to 0.07) (Supplementary Figure 3B, E).

When analyses were restricted to individuals with early-onset disease, the overall limbic-predominant pattern was preserved, but effect estimates were generally less precise. LATE-NC was associated with a trend toward smaller amygdala (β = −40.0 mm^3^, 95% CI −102.9 to 17.7; AUC = 0.72, 95% CI 0.57 to 0.85) and hippocampal volumes (β = −141.1 mm^3^, 95% CI −319.5 to 21.9; AUC = 0.74, 95% CI 0.58 to 0.89) (Supplementary Figure 5A-B). Regional cortical analyses similarly showed a directionally consistent pattern of lower gray matter volumes in the bilateral entorhinal (left: β = −25.8 mm^3^, 95% CI −115.4 to 53.9; right: β = −32.3 mm^3^, 95% CI −103.4 to 36.4) and medial orbitofrontal cortices (left: β = −105 mm^3^, 95% CI −259.6 to 51; right: β = −96.0 mm^3^, 95% CI −230.3 to 41.3) (Supplementary Figure 5C). By contrast, little evidence of group differences was observed for the ERC/PHC ratio (β = −0.00, 95% CI −0.06 to 0.06; AUC = 0.56, 95% CI 0.43 to 0.72) (Supplementary Figure 3C).

### LATE-NC is associated with a similar limbic-predominant pattern of hypometabolism, whereas previously proposed IMT ratio shows limited discriminative power in early-onset cases

Among the 71 participants with FDG PET, LATE-NC was also associated with a pattern of lower uptake in the bilateral entorhinal (left: β = −0.067, 95% CI −0.129 to 0.000; right: β = - 0.084, 95% CI −0.155 to −0.015) and medial orbitofrontal cortices (left: β = −0.055, 95% CI - 0.134 to 0.023; right: β = −0.071, 95% CI −0.136 to −0.004), adjusting for age at symptom onset (Figure 5A). In keeping with these findings, the previously proposed IMT ratio tended to be higher in individuals with LATE-NC (β = 0.064, 95% CI −0.001 to 0.135; AUC = 0.68, 95% CI 0.50 to 0.82), with greater separation observed at later ages at symptom onset (Figure 5B). In interaction analyses, every 10-year increase in age at symptom onset was associated with a higher IMT ratio (β = 0.068, 95% CI 0.025 to 0.138) among individuals with LATE-NC, with no corresponding association among those without (Figure 5C).

**Figure 5.**
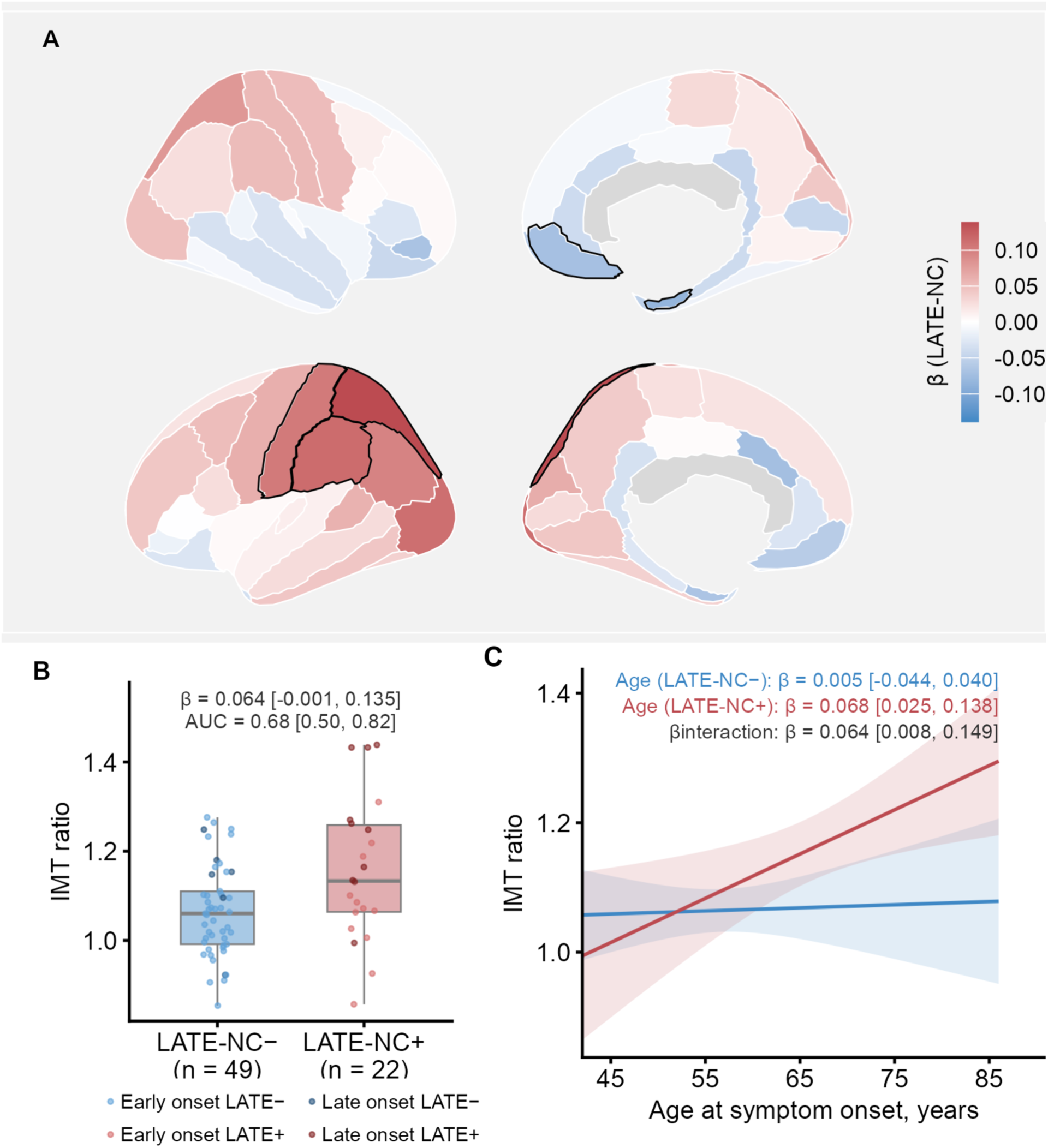
Regional FDG-PET metabolism according to LATE-NC status and age at symptom onset. (A) Cortical metabolic differences associated with LATE-NC (LATE-NC+ vs. LATE-NC-). Upper and lower views correspond to the right and left hemispheres, respectively. Dark outlines mark regions where the 95% CIs did not include zero. (B) Inferior-to-medial temporal (IMT) ratio by LATE-NC status. Boxplot shows the median and interquartile range, with individual observations stratified by early- and late-onset cases. β coefficients (95% CI) correspond to the association between LATE-NC and IMT ratio; AUC value reflects the discriminative performance of the IMT ratio for LATE-NC status. (C) Model-estimated IMT ratio across age at symptom onset by LATE-NC status. Lines represent model estimates and shaded areas represent 95% CIs. Models were adjusted for age at symptom onset, with 95% CIs estimated using 5,000 resamples. AUC, area under the curve; CI, confidence interval; FDG-PET, ^18^F-fluorodeoxyglucose positron emission tomography; IMT, inferior-to-medial temporal; LATE-NC, limbic-predominant age-related TDP-43 encephalopathy neuropathologic change.

In sensitivity analyses excluding individuals with LATE-NC stage 1, the regional pattern of cortical hypometabolism remained directionally consistent with the primary analysis, with trends toward lower metabolism in the entorhinal (left: β = −0.067, 95% CI −0.137 to 0.001; right: β = −0.081, 95% CI −0.165 to 0.003) and medial orbitofrontal (left: β = −0.027, 95% CI - 0.099 to 0.046; right: β = −0.04, 95% CI −0.108 to 0.03) cortices in the LATE-NC stage 2-3 group (Supplementary Figure 6A). The IMT ratio was higher in this group (β = 0.091, 95% CI 0.018 to 0.172; AUC = 0.75, 95% CI 0.59 to 0.88) and remained positively associated with age at symptom onset (β = 0.046 per 10-year increase, 95% CI 0.004 to 0.117) (Supplementary Figure 6B-C).

When analyses were restricted to individuals with early-onset disease, a similar limbic-predominant pattern of hypometabolism was observed in the bilateral entorhinal (left: β = - 0.069, 95% CI −0.138 to 0.005; right: β = −0.089, 95% CI −0.153 to −0.025) and medial orbitofrontal (left: β = −0.086, 95% CI −0.174 to 0.002; right: β = −0.109, 95% CI −0.182 to − 0.037) cortices (Supplementary Figure 6D). However, the IMT ratio showed little evidence of separation between groups (β = 0.034, 95% CI −0.038 to 0.111; AUC = 0.59, 95% CI 0.44 to 0.76) (Supplementary Figure 6E).

### LATE-NC shows no evidence of association with amyloid or medial temporal tau PET measures

Among the 75 participants with amyloid PET, global amyloid burden (Centiloids) did not differ by LATE-NC status (β = 0.70, 95% CI −21.50 to 19.17; AUC = 0.55, 95% CI 0.45 to 0.67) after adjusting for age at symptom onset. There was also no evidence of an interaction between LATE-NC status and age at symptom onset (β interaction = 6.25 per 10-year increase, 95% CI −16.27 to 23.88) (Supplementary Figure 7A-B). In exploratory analyses of the 45 participants with tau PET, medial temporal tau PET uptake (amygdala, entorhinal and parahippocampal cortices) likewise did not differ by LATE-NC status (β = 0.055, 95% CI −0.126 to 0.243; AUC = 0.61, 95% CI 0.45 to 0.79), with no evidence of an interaction with age at symptom onset (β interaction = −0.009 per 10-year increase, 95% CI −0.242 to 0.174) (Supplementary Figure 7C-D).

### LATE-NC shows no evidence of association with longitudinal clinical progression

In linear mixed-effects models adjusted for age at symptom onset, with time measured from the first available assessment for each outcome, rates of change in MMSE (LATE-NC × time interaction: β = 0.03, 95% CI −0.93 to 0.98 points/year) or CDR-SB (LATE-NC × time interaction: β = 0.31, 95% CI −0.18 to 0.80 points/year) did not differ significantly by LATE-NC status (Figure 6A-B). In MRI models, estimates pointed toward faster amygdala (LATE-NC × time interaction: β = −11.3, 95% CI −25.8 to 3.2 mm^3^/year) and hippocampal atrophy (LATE-NC × time interaction: β = −27.65, 95% CI −81.77 to 26.48 mm^3^/year) with LATE-NC, although neither LATE-NC × time interaction reached statistical significance (Figure 6C-D).

**Figure 6.**
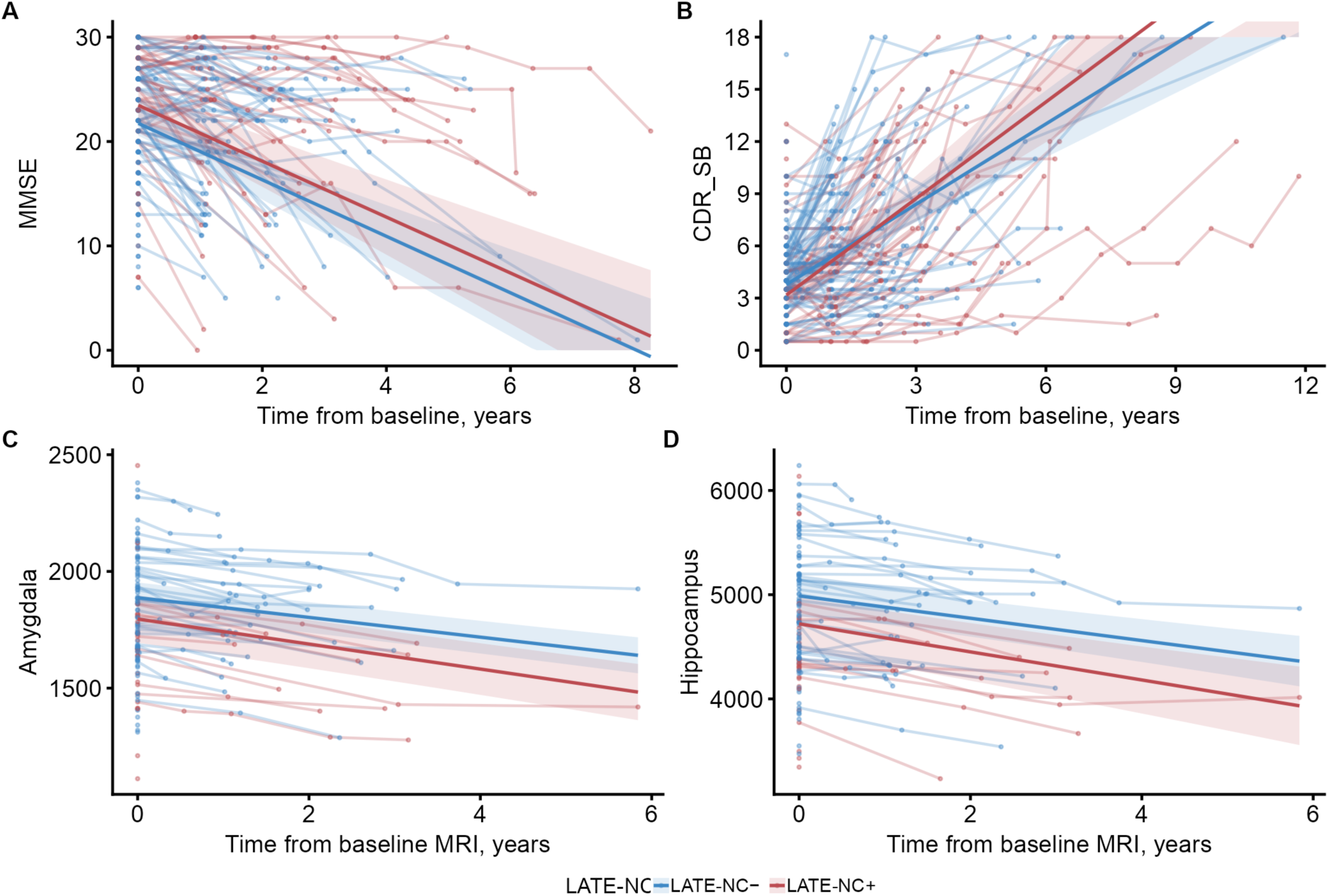
Longitudinal clinical and structural MRI progression according to LATE-NC status. Model-estimated longitudinal changes of (A) MMSE, (B) CDR-SB, (C) amygdala gray matter volume, and (D) hippocampal gray matter volume according to LATE-NC status. Lines represent model estimates and shaded areas represent 95% CIs. Individual observations and within-participant changes are shown in lighter colors. Clinical models were adjusted for age at symptom onset, with sex and education retained when they improved model fit, and structural MRI models were adjusted for age at symptom onset and total intracranial volume. CDR-SB, Clinical Dementia Rating Sum of Boxes; CI, confidence interval; LATE-NC, limbic-predominant age-related TDP-43 encephalopathy neuropathologic change; MMSE, Mini-Mental State Examination.

## Discussion

In this neuropathologically confirmed sample of clinically impaired individuals with AD enriched for early age at symptom onset (mean age = 62.6 ± 10.5 years), we identified a significant frequency of co-occurring LATE-NC (31%). Importantly, although LATE-NC retained its predominantly limbic signature at the group level, selected clinical and neuroimaging features were more pronounced with older age at symptom onset within the LATE-NC group, particularly for presenting symptoms and neuroimaging markers that may reflect the combined contribution of AD and LATE-NC pathology. The convergence across clinical, imaging measures and sensitivity analyses supports the robustness of the overall pattern.

Aging and ADNC are the two factors most consistently associated with the presence of LATE-NC.^3–5^ Individuals with high ADNC are approximately 2-2.5 times more likely to harbor concomitant LATE-NC than those lacking ADNC.^3^ This robust association has led to the hypothesis that medial temporal tau pathology promotes TDP-43 aggregation, consistent with the frequent colocalization of both pathologies within vulnerable hippocampal neurons.^3,24–26^ However, this seeding model alone does not fully explain the consistently lower prevalence of LATE-NC across younger AD populations. Despite comparable medial temporal tau burden^21,22,27^, LATE-NC prevalence in EOAD, ADAD and DSAD remains markedly lower than in late-onset AD^18–20,22,28^. This pattern was replicated in the present study, where LATE-NC prevalence increased with age at symptom onset despite no detectable difference in ADNC severity or medial temporal tau PET uptake by LATE-NC status. These observations suggest that age-related biological susceptibility may contribute to LATE-NC occurrence. Emerging genetic evidence supports a role for biological susceptibility, as established LATE-NC risk genes converge on pathways governing lipid metabolism, lysosomal function, and cellular homeostasis, processes that become increasingly vulnerable with aging.^2,29–31^ Consistent with this, our cohort showed that individuals carrying multiple genetic risk factors, such as *APOE* ε4 carrier status and the *TMEM106B* rs1990622 risk genotype, were more frequently represented among those with LATE-NC, including among early-onset cases.

While LATE-NC is consistently associated with a limbic-predominant signature in AD,^3,4,6–8^ our findings suggest that its clinical and neuroimaging expression is also shaped by the biological characteristics of the underlying AD. In this study, several features of the LATE-NC phenotype remained evident across the age-at-onset continuum, including a higher frequency of an amnestic clinical phenotype, greater episodic memory impairment, and greater medial temporal involvement on structural MRI and FDG PET. However, other features showed a clearer age dependence: the likelihood of memory being the presenting symptom and smaller amygdala and hippocampal volumes became more apparent with older age at symptom onset. This age dependence was particularly evident in the IMT metabolism ratio on FDG PET, a measure of relative regional vulnerability. These findings should be interpreted in the context of the distinctive features of EOAD. Compared with late-onset cases, EOAD more commonly presents with multidomain cognitive impairment and greater involvement of non-amnestic domains, whereas a predominantly amnestic single-domain presentation is relatively uncommon.^32^ This clinical heterogeneity is accompanied by a more widespread distribution of tau pathology, with greater burden across neocortical regions, while differences in medial temporal tau burden are comparatively less pronounced.^22,27^ A similar predominance of neocortical involvement is observed for cortical atrophy, particularly in frontoparietal and posterior lateral temporal regions, with relative preservation of the medial temporal lobe compared with late-onset AD.^32–34^ Thus, the medial temporal-predominant signature observed in individuals with concomitant LATE-NC may reflect the contribution of LATE-NC, an underlying pattern of AD with greater medial temporal vulnerability, or the combined effect of both pathologies. In younger-onset AD, greater neocortical involvement may reduce the relative predominance of medial temporal impairment, particularly for measures that rely on relative medial temporal versus neocortical involvement. More broadly, these findings extend the growing recognition that the clinical manifestation of AD reflects the convergence of multiple concomitant pathologies,^2,12,19,21,35^ suggesting that the clinical and neuroimaging expression of LATE-NC should be interpreted in the context of the underlying AD.

The current *in vivo* frameworks for identifying concomitant LATE-NC represent an important step toward the recognition of TDP-43 pathology during life in the absence of molecular biomarkers. These approaches combine characteristic clinical and neuroimaging features of LATE-NC with measures of disproportionate clinical impairment relative to tau burden (referred to as tau-clinical mismatch) to identify individuals most likely to harbor concomitant TDP-43 pathology.^15,17,35^ However, in this cohort, selected clinical and neuroimaging features currently used to infer LATE-NC became less pronounced at younger age at symptom onset, suggesting that their interpretation may require additional consideration when applied to younger AD populations. Moreover, the greater AD tau burden characteristic of EOAD may also limit the applicability of tau-clinical mismatch approaches.^21,27^ In our cohort, LATE-NC occurred despite comparable ADNC levels, and our limited, exploratory tau PET analyses did not detect differences in medial temporal tau burden by LATE-NC status. Longitudinally, our analyses did not identify a clear association between LATE-NC and clinical progression, despite a longer overall disease duration among individuals with the co-pathology. This is consistent with previous neuropathological observations in younger cohorts with a similar degree of baseline global cognitive impairment, where the association of LATE-NC with cognitive decline or progression to dementia was less evident.^5,22^ The greater baseline cognitive and functional impairment in some cohorts reporting accelerated decline with concomitant AD and LATE-NC raises the possibility that its clinical impact may vary across the disease course.^3,6–9,12^ The distinct clinical phenotype and disease trajectory of younger-onset AD may additionally influence the ability to detect an independent association of LATE-NC with progression. These observations are particularly relevant in the era of disease-modifying therapies, in which identifying concomitant LATE-NC has become increasingly important for refining prognosis, patient stratification, and interpreting therapeutic response.^2,4,35^

The strengths of this study include one of the largest neuropathologically confirmed cohorts enriched for EOAD examined for LATE-NC to date. Whereas previous reports of LATE-NC in younger-onset AD were largely limited to post-mortem characterization, combining autopsy confirmation with in vivo clinical, cognitive, multimodal neuroimaging, genetic and longitudinal data allowed us to characterize its clinical and neuroimaging expression across the AD age-at-onset continuum. Nevertheless, some methodological limitations should be considered when interpreting these findings. As is standard practice across brain banks, neuropathological assessment was based predominantly on unilateral brain sampling, which may have resulted in underdetection of LATE-NC or underestimation of its pathological stage. The number of early-onset individuals with LATE-NC remained limited because of the relatively low frequency of the pathology in younger populations. Sample sizes also differed across imaging modalities, with smaller sample sizes for PET imaging, which reduced statistical power in these analyses. Thus, imaging findings should be interpreted within the overall pattern of evidence across modalities. Longitudinal analyses were further limited by the relatively small number of participants with repeated assessments and the potential for a floor effect in domain-specific neuropsychological measures. Consequently, longitudinal analyses were restricted to global cognitive and functional measures. Finally, although the determination of age at symptom onset is inherently susceptible to bias, it represents a clinically relevant estimate of disease onset and is less influenced by healthcare access and by differences in disease aggressiveness that may affect the timing of clinical assessment and death.

Taken together, our findings suggest that older age at symptom onset is associated with more frequent LATE-NC in AD and, when present, more overt limbic-predominant expression. However, concomitant LATE-NC should still be considered in EOAD with prominent episodic memory impairment, even when other features typically associated with LATE-NC in older populations are less evident. Studying EOAD alongside later-onset AD offers a model for exploring the relative contributions of AD pathology, aging and biological susceptibility to LATE-NC. Further studies incorporating sporadic EOAD, ADAD and DSAD alongside late-onset AD and LATE-NC without significant AD neuropathology may clarify the biological drivers of LATE-NC, inform *in vivo* biomarker development and refine disease stratification, prognosis, and interpretation of treatment response across the AD spectrum.

## Methods

### Participants

We queried the UCSF NDBB database to identify individuals with intermediate-to-high ADNC (score 2-3), according to National Institute on Aging-Alzheimer’s Association (NIA-AA) criteria for the neuropathological assessment of AD.^36^ Cases with FTLD-TDP or FTLD-tau were excluded. A total of 223 cases met the inclusion criteria (Figure 1). Eighteen cases were excluded because of incomplete LATE-NC staging, and a further nine were excluded because they lacked cognitive or functional impairment, defined as a Clinical Dementia Rating (CDR) global score of 0. The final study cohort comprised 196 individuals.

All participants or their legally authorized representatives provided informed consent for brain donation and research participation. The study was approved by the UCSF Institutional Review Board.

### Neuropathological assessment

Neuropathological evaluations were performed by experts in the neuropathology of dementia (S.S., D.L.F., W.W.S., L.T.G.) between April 2018 and October 2025. Three cases were evaluated elsewhere, and diagnostic conclusions were verified by the UCSF team. Post-mortem brain tissue was processed according to standardized NDBB protocols. Twenty-four predefined anatomical regions were systematically sampled and evaluated following established research procedures, and histological and immunohistochemical methods have been described previously.^28^

Intermediate-to-high ADNC was defined using the ABC scoring system, applying standardized scoring of Thal amyloid-β phase, Braak neurofibrillary tangle stage, and Consortium to Establish a Registry for AD (CERAD) scale for neuritic plaque density.^36–39^ LATE-NC was assessed using TDP-43 immunohistochemistry.^40^ All cases were systematically examined for TDP-43 proteinopathy in the amygdala, hippocampal formation, anterior cingulate gyrus, inferior temporal gyrus, and middle frontal gyrus. LATE-NC was staged according to current consensus recommendations based on the hierarchical anatomical distribution of TDP-43 pathology involving the amygdala (stage 1), hippocampus/entorhinal cortex (stage 2), and middle frontal gyrus (stage 3).^4^ Additional co-pathologies, including Lewy body disease, cerebral amyloid angiopathy, hippocampal sclerosis, and vascular brain injury, were assessed according to established neuropathological criteria.^41–44^

### Clinical assessment

Of the 196 participants, 181 had been enrolled during life in longitudinal research studies through the UCSF Alzheimer’s Disease Research Center (ADRC) at the Edward and Pearl Fein Memory and Aging Center, where prospectively collected data included standardized clinical evaluations including medical history, neurological examination, neuropsychological assessment, and, when available, neuroimaging and genetic testing. Clinical information for the remaining 15 participants was obtained through review of available clinical records as part of the UCSF NDBB neuropathological assessment.

Clinical variables included age at symptom onset, sex, years of education, age at death, disease duration, presenting symptom, and clinical phenotype. Age at symptom onset was recorded in the database for 193 participants and was defined as the age at which the first cognitive or behavioral symptom attributable to neurodegenerative disease was judged to have occurred based on the participant’s clinical history, informant interview, and/or initial clinical evaluation at UCSF. The three remaining participants were classified as early- or late-onset based on clinical records. The presenting symptom was defined as the predominant cognitive or behavioral symptom reported at disease onset. Disease duration was calculated as the interval between symptom onset and death. Diagnoses were established at multidisciplinary consensus conferences led by expert behavioral neurologists, and clinical phenotype was classified according to the predominant clinical syndrome as amnestic, primary progressive aphasia (PPA), posterior cortical atrophy (PCA), or other non-amnestic presentations, including dysexecutive and other atypical presentations not classified as PPA or PCA.^45–47^

The standardized neuropsychological battery and scoring procedures have been described previously.^48^ Representative measures of each cognitive domain were selected, including CVLT and Benson Figure 10-minute delayed recall for episodic memory, Modified Trails and Digit Span Backward (maximum span) for executive function, Animal Fluency and the 15-item Boston Naming Test for language, Visual Object and Space Perception (VOSP) Number Location for visuospatial abilities, and Calculation. Longitudinal cognitive and functional progression were evaluated using repeated MMSE scores from 165 participants (395 observations; mean 2.4 observations per participant) and CDR-SB scores from 180 participants (386 observations; mean 2.1 observations per participant).

### Genetic analyses

Genetic screening for pathogenic variants in *APP*, *PSEN1*, *PSEN2*, *MAPT*, *GRN*, *TARDBP*, and *FUS* (n = 169), *C9orf72* expansion testing (n = 174), *APOE* genotyping (n = 175) and *TMEM106B* rs1990622 genotyping (n = 171) were performed. Genetic testing procedures have been described previously.^49,50^ For *TMEM106B* rs1990622, genomic DNA was extracted from peripheral blood using standard protocols and genotyped using a commercially available TaqMan® SNP assay (C_11171598_20; Thermo Fisher Scientific) on LightCycler® 480 or QuantStudio™ real-time PCR system. *TMEM106B* rs1990622 was classified as A/A, A/G or G/G genotype. Assays were run in duplicate. *APOE* was analyzed according to ε4 carrier status.

### Neuroimaging acquisition and processing

Structural MRI, FDG PET, amyloid PET, and tau PET acquisition protocols, imaging preprocessing, and regional quantification procedures have been described previously.^51^

#### Structural MRI

Structural MRI was available for 116 participants (192 scans including longitudinal follow-up). MRI scans were performed at the UCSF Neuroimaging Center using 3T Siemens scanners, including Trio Tim (76 baseline scans; 113 scans overall) and Prisma Fit (40 baseline scans; 79 scans overall), with standardized T1-weighted magnetization-prepared rapid gradient echo (MPRAGE) acquisition protocols (1.0 mm isotropic resolution; TR = 2300 ms; TE = 2.9-2.98 ms; TI = 900 ms; flip angle = 9°). T1-weighted images underwent visual quality control, N3 bias field correction,^52^ and tissue segmentation using Statistical Parametric Mapping version 12 (SPM12, https://www.fil.ion.ucl.ac.uk/spm/) unified segmentation.^53^ Customized group template was generated using the *Large Deformation Diffeomorphic Metric Mapping* framework, followed by spatial normalization, modulation, and smoothing with an 8-mm full-width half-maximum Gaussian kernel. Regional gray matter volumes were derived according to the Desikan-Killiany atlas, and TIV was obtained for volumetric analyses. ERC/PHC ratio was calculated as the mean bilateral entorhinal gray matter volume divided by the mean bilateral parahippocampal gray matter volume.

#### PET imaging

FDG PET was available for 71 participants, with scans acquired using Siemens Biograph 6 TruePoint PET/CT (n = 52) or Siemens ECAT EXACT HR (n = 19) scanners during a 30-60 min post-injection window (∼10 mCi). Amyloid PET was available for 75 participants, including 65 [^11^C]Pittsburgh compound-B (PiB), six [^18^F]florbetaben and four [^18^F]florbetapir scans. Amyloid PET scans were acquired using Siemens Biograph 6 TruePoint PET/CT (n = 44), Siemens ECAT EXACT HR (n = 21), GE Discovery STE PET/CT (n = 7), or Siemens Biograph Vision PET/CT (n = 3) scanners during 50-70 min post-injection for PiB (∼10-15 mCi) and florbetapir (∼10 mCi) and 90-110 min post-injection for florbetaben (∼8 mCi). Tau PET with [^18^F]flortaucipir was available for 45 participants, with scans acquired using Siemens Biograph 6 TruePoint PET/CT (n = 41) or GE Discovery STE PET/CT (n = 4) scanners. Tau PET scans were acquired during an 80-100 min post-injection window following the intravenous administration of the tracer (∼10 mCi).

All PET images were coregistered to each participant’s T1-weighted MRI, and voxel-wise standardized uptake value ratio (SUVR) images were generated in native MRI space using tracer-specific reference regions: pons for FDG PET,^54^ whole cerebellum for amyloid PET^55^, and inferior cerebellar gray matter for tau PET.^56^ Smoothing with a differential kernel was applied to harmonize spatial resolution. Automated anatomical segmentation was performed in native MRI space using FreeSurfer (version 7, https://surfer.nmr.mgh.harvard.edu/fswiki), with regions of interest (ROIs) defined according to the Desikan-Killiany atlas, from which regional SUVR values were subsequently extracted. Additionally, IMT ratio was calculated based on FDG PET by dividing inferior temporal cortex SUVR by a voxel-weighted mean SUVR of the amygdala and hippocampus. To quantify global cortical amyloid burden, amyloid PET SUVR in composite cortical region was converted to Centiloid values using tracer-and pipeline-specific conversion equations.^55,57^ Medial temporal tau burden was calculated as a voxel-weighted mean flortaucipir SUVR across the bilateral amygdala, entorhinal cortex, and parahippocampal cortex.

### Statistical analysis

Continuous variables are presented as mean and standard deviation, and categorical variables as counts and percentages. The distribution of continuous variables was assessed using the Shapiro-Wilk test, and homogeneity of variance was evaluated using Levene’s test. Baseline demographic and clinical variables were compared between participants with and without LATE-NC using Student’s t-tests for continuous variables and χ^2^ tests for categorical variables. Differences in genotype frequencies were assessed using Fisher’s exact test.

Cross-sectional associations between LATE-NC and clinical, cognitive, and neuroimaging outcomes were evaluated using multiple regression models. Logistic regression models were used for binary outcomes, whereas linear regression models were performed for continuous outcomes. Models evaluating clinical outcomes were adjusted for age at symptom onset, sex, and years of education, with covariates retained when they improved model fit. Structural MRI models were adjusted for age at symptom onset and TIV; sex was excluded as it did not improve model fit in most regions. PET models were only adjusted for age at symptom onset because of the limited sample size. For medial temporal regional measures and derived ratios on structural MRI and FDG PET, receiver operating characteristic (ROC) analyses were additionally performed to assess discrimination between individuals with and without LATE-NC, quantified using AUC. Interactions between LATE-NC and age at symptom onset were evaluated to determine whether the effects of LATE-NC varied across the age-at-onset continuum. Bootstrap resampling with 5,000 iterations was applied to all cross-sectional regression models (seed = 1234).

Longitudinal progression of cognitive, functional, and structural MRI outcomes was evaluated using linear mixed-effects models. Fixed effects included time from baseline (first available value), LATE-NC status, age at symptom onset, and the interactions between time and LATE-NC and between time and age at symptom onset. For clinical outcomes, sex and years of education were additionally included when they improved model fit, whereas longitudinal structural MRI models were adjusted for TIV. Alternative covariance structures and random-effects structures (random intercepts and random slopes) were compared using Akaike’s Information Criterion (AIC) and Bayesian Information Criterion (BIC), and the most parsimonious model was selected for each outcome.

Two sets of sensitivity analyses were conducted. First, given that LATE-NC stage 1 has been proposed to represent an early pathological stage with limited clinical expression,^3^ analyses were repeated after excluding individuals with LATE-NC stage 1 (i.e. stage 0 versus stages 2-3 combined). Additional analyses restricted to individuals with EOAD (symptom onset at or before 65 years of age) were performed to assess the robustness of the findings independent of the higher frequency of LATE-NC with increasing age at symptom onset.

All statistical tests were two-sided, with statistical significance defined as P < 0.05. Regression estimates are reported as odds ratios (ORs) or regression coefficients (β) with corresponding 95% confidence intervals (CIs).

Statistical analyses were performed using R version 4.5.0 (R Foundation for Statistical Computing, Vienna, Austria). Data management and preprocessing were performed using the *dplyr* (v1.2.1), *tidyr* (v1.3.1) and *skimr* (v2.2.1) packages. Statistical analyses were conducted using the *MASS* (v7.3.65), *nlme* (v3.1.168) and *boot* (v1.3.32) packages, and figures were generated using *ggplot2* (v4.0.0) and *ggseg* (v2.1.1).

## Data availability

Supplementary Information is provided with this paper. Deidentified data supporting the findings of this study are available to qualified academic investigators upon reasonable request through the UCSF Edward and Pearl Fein Memory and Aging Center (memory.ucsf.edu/research-trials/professional/open-science), subject to institutional approvals and applicable data-sharing policies.

## Code availability

The custom R script used to perform the analyses in this study is available from the corresponding author upon reasonable request.

## Acknowledgements

Samples from the National Centralized Repository for Alzheimer’s Disease and Related Dementias (NCRAD), which receives government support under a cooperative agreement grant (U24 AG021886) awarded by the National Institute on Aging (NIA), were used in this study. We thank contributors who collected samples used in this study, as well as patients and their families, whose help and participation made this work possible.

Large language models were used for language editing. All content was reviewed and verified by the authors, who take full responsibility for the final manuscript.

## Author contributions

Conceptualization: N.Z., R.L.J. and G.D.R. Methodology: N.Z., R.L.J. and G.D.R. Formal analysis: N.Z. Data curation: N.Z., R.L.J. and S.S. Writing – original draft: N.Z., R.L.J. and G.D.R. Writing – review and editing: all authors. Supervision: R.L.J. and G.D.R. N.Z. and R.L.J. accessed and verified the underlying data. I.E.A. reviewed the statistical methodology and analytical approach. All authors contributed to the interpretation of the results, critically revised the manuscript, approved the final version, and accept responsibility for the decision to submit the manuscript.

## References

1. Coulthard EJ, Love S. A broader view of dementia: multiple co-pathologies are the norm. Brain. 2018;141(7):1894–1897. doi:10.1093/brain/awy153

2. Robinson JL, Lee EB, Xie SX, et al. Neurodegenerative disease concomitant proteinopathies are prevalent, age-related and APOE4-associated. Brain. 2018;141(7):2181–2193. doi:10.1093/brain/awy146

3. Nelson PT, Brayne C, Flanagan ME, et al. Frequency of LATE neuropathologic change across the spectrum of Alzheimer’s disease neuropathology: combined data from 13 community-based or population-based autopsy cohorts. Acta Neuropathol. 2022;144(1):27–44. doi:10.1007/s00401-022-02444-1

4. Nelson PT, Dickson DW, Trojanowski JQ, et al. Limbic-predominant age-related TDP-43 encephalopathy (LATE): consensus working group report. Brain. 2019;142(6):1503–1527. doi:10.1093/brain/awz099

5. Wang SHJ, Guo Y, Ervin JF, Lusk JB, Luo S. Neuropathological associations of limbic-predominant age-related TDP-43 encephalopathy neuropathological change (LATE-NC) differ between the oldest-old and younger-old. Acta Neuropathol. 2022;144(1):45–57. doi:10.1007/s00401-022-02432-5

6. Josephs KA, Dickson DW, Tosakulwong N, et al. Rates of hippocampal atrophy and presence of post-mortem TDP-43 in patients with Alzheimer’s disease: a longitudinal retrospective study. Lancet Neurol. 2017;16(11):917–924. doi:10.1016/S1474-4422(17)30284-3

7. Kapasi A, Yu L, Boyle PA, Barnes LL, Bennett DA, Schneider JA. Limbic-predominant age-related TDP-43 encephalopathy, ADNC pathology, and cognitive decline in aging. Neurology. 2020;95(14):e1951–e1962. doi:10.1212/WNL.0000000000010454

8. Josephs KA, Whitwell JL, Weigand SD, et al. TDP-43 is a key player in the clinical features associated with Alzheimer’s disease. Acta Neuropathol. 2014;127(6):811–824. doi:10.1007/s00401-014-1269-z

9. Thomas DX, Bajaj S, McRae-McKee K, Hadjichrysanthou C, Anderson RM, Collinge J. Association of TDP-43 proteinopathy, cerebral amyloid angiopathy, and Lewy bodies with cognitive impairment in individuals with or without Alzheimer’s disease neuropathology. Sci Rep. 2020;10(1):14579. doi:10.1038/s41598-020-71305-2

10. Boyle PA, Yu L, Leurgans SE, et al. Attributable risk of Alzheimer’s dementia attributed to age-related neuropathologies. Ann Neurol. 2019;85(1):114–124. doi:10.1002/ana.25380

11. De Flores R, Wisse LEM, Das SR, et al. Contribution of mixed pathology to medial temporal lobe atrophy in Alzheimer’s disease. Alzheimers Dement. 2020;16(6):843–852. doi:10.1002/alz.12079

12. Ngam PI, Anzai Y, Cliatt Brown CJ, et al. Copathologies of Limbic-Predominant Age-Related TDP-43 Encephalopathy and Alzheimer Disease: [18F]FDG PET Statistical Mapping and Quantitative MRI Volumetry. J Nucl Med. 2026;67(4):621–628. doi:10.2967/jnumed.125.270614

13. Grothe MJ, Moscoso A, Silva-Rodríguez J, et al. Differential diagnosis of amnestic dementia patients based on an FDG-PET signature of autopsy-confirmed LATE-NC. Alzheimers Dement. 2023;19(4):1234–1244. doi:10.1002/alz.12763

14. Wisse LEM, Wuestefeld A, Murray ME, Jagust W, La Joie R. Role of tau versus TDP-43 pathology on medial temporal lobe atrophy in aging and Alzheimer’s disease. Alzheimers Dement. 2025;21(2):e14582. doi:10.1002/alz.14582

15. Groot C, Calandri IL, Bader I, et al. Characterizing Individuals Fulfilling Clinical Criteria for Limbic-Predominant Age-Related TDP-43 Encephalopathy in a Tertiary Memory Clinic. Neurology. 2026;106(9):e214810. doi:10.1212/WNL.0000000000214810

16. Buciuc M, Botha H, Murray ME, et al. Utility of FDG-PET in diagnosis of Alzheimer-related TDP-43 proteinopathy. Neurology. 2020;95(1):e23–e34. doi:10.1212/WNL.0000000000009722

17. Wolk DA, Nelson PT, Apostolova L, et al. Clinical criteria for limbic-predominant age-related TDP-43 encephalopathy. Alzheimers Dement. 2025;21(1):e14202. doi:10.1002/alz.14202

18. Davidson YS, Raby S, Foulds PG, et al. TDP-43 pathological changes in early onset familial and sporadic Alzheimer’s disease, late onset Alzheimer’s disease and Down’s Syndrome: association with age, hippocampal sclerosis and clinical phenotype. Acta Neuropathol. 2011;122(6):703–713. doi:10.1007/s00401-011-0879-y

19. Flores-Aguilar L, Zaikos TD, Rivera I, et al. Frequency of mixed neuropathologies in individuals with Down syndrome with and without Alzheimer’s dementia. Acta Neuropathol. 2026;151(1):55. doi:10.1007/s00401-026-03028-z

20. Ichimata S, Yoshida K, Visanji NP, Lang AE, Nishida N, Kovacs GG. Patterns of Mixed Pathologies in Down Syndrome. J Alzheimers Dis. 2022;87(2):595–607. doi:10.3233/JAD-215675

21. Spina S, La Joie R, Petersen C, et al. Comorbid neuropathological diagnoses in early versus late-onset Alzheimer’s disease. Brain. 2021;144(7):2186–2198. doi:10.1093/brain/awab099

22. Smirnov DS, Salmon DP, Galasko D, et al. Association of Neurofibrillary Tangle Distribution With Age at Onset–Related Clinical Heterogeneity in Alzheimer Disease: An Autopsy Study. Neurology. 2022;98(5). doi:10.1212/WNL.0000000000013107

23. Wegiel J, Flory M, Kuchna I, et al. Developmental deficits and staging of dynamics of age associated Alzheimer’s disease neurodegeneration and neuronal loss in subjects with Down syndrome. Acta Neuropathol Commun. 2022;10(1):2. doi:10.1186/s40478-021-01300-9

24. Shahidehpour RK, Nelson PT, Srinivasan S, Yu Z, Bachstetter AD. Assessing Co-Localization of ITM2B With Alzheimer’s Disease and Limbic-Predominant Age-Related TDP-43 Encephalopathy Neuropathologic Changes. Neuropathology. 2025;45(4):e70003. doi:10.1111/neup.70003

25. Josephs KA, Murray ME, Tosakulwong N, et al. Pathological, imaging and genetic characteristics support the existence of distinct TDP-43 types in non-FTLD brains. Acta Neuropathol. 2019;137(2):227–238. doi:10.1007/s00401-018-1951-7

26. Smith VD, Bachstetter AD, Ighodaro E, et al. Overlapping but distinct TDP-43 and tau pathologic patterns in aged hippocampi. Brain Pathol. 2018;28(2):264–273. doi:10.1111/bpa.12505

27. Chiotis K, Blazhenets G, Soleimani-Meigooni DN, et al. Distinct Tau PET Dynamics in Early vs. Late Age-of-Onset Alzheimer’s disease. medRxiv. Preprint posted online January 26, 2026. doi:10.64898/2026.01.25.26344738

28. Spina S, Brown JA, Deng J, et al. Neuropathological correlates of structural and functional imaging biomarkers in 4-repeat tauopathies. Brain. 2019;142(7):2068–2081. doi:10.1093/brain/awz122

29. Acewicz A, Tarka S, Grzegorczyk M, Rzepliński R, Wierzba-Bobrowicz T, Stępień T. Associations between TMEM106B C-terminal fragment aggregation, age, and TDP-43 or tau pathology. Brain Pathol. 2026;36(5):e70090. doi:10.1111/bpa.70090

30. Meneses A, Koga S, O’Leary J, Dickson DW, Bu G, Zhao N. TDP-43 Pathology in Alzheimer’s Disease. Mol Neurodegener. 2021;16(1):84. doi:10.1186/s13024-021-00503-x

31. Nelson PT, Fardo DW, Wu X, Aung KZ, Cykowski MD, Katsumata Y. Limbic-predominant age-related TDP-43 encephalopathy (LATE-NC): Co-pathologies and genetic risk factors provide clues about pathogenesis. J Neuropathol Exp Neurol. 2024;83(6):396–415. doi:10.1093/jnen/nlae032

32. Putcha D, Katsumi Y, Touroutoglou A, et al. Heterogeneous clinical phenotypes of sporadic early-onset Alzheimer’s disease: a neuropsychological data-driven approach. Alzheimers Res Ther. 2025;17(1):38. doi:10.1186/s13195-025-01689-8

33. Touroutoglou A, Katsumi Y, Brickhouse M, et al. The Sporadic Early-onset Alzheimer’s Disease Signature Of Atrophy: Preliminary Findings From The Longitudinal Early-onset Alzheimer’s Disease Study (LEADS) Cohort. Alzheimers Dement. 2023;19(S9):S74–S88. doi:10.1002/alz.13466

34. Migliaccio R, Agosta F, Possin KL, et al. Mapping the Progression of Atrophy in Early- and Late-Onset Alzheimer’s Disease. J Alzheimers Dis. 2015;46(2):351–364. doi:10.3233/JAD-142292

35. Brown CA, Mundada NS, Cousins KAQ, et al. Evaluation of Copathology and Clinical Trajectories in Individuals With Tau-Clinical Mismatch. JAMA Neurol. 2026;83(2):126–136. doi:10.1001/jamaneurol.2025.4974

36. Montine TJ, Phelps CH, Beach TG, et al. National Institute on Aging–Alzheimer’s Association guidelines for the neuropathologic assessment of Alzheimer’s disease: a practical approach. Acta Neuropathol. 2012;123(1):1–11. doi:10.1007/s00401-011-0910-3

37. Braak H, Alafuzoff I, Arzberger T, Kretzschmar H, Del Tredici K. Staging of Alzheimer disease-associated neurofibrillary pathology using paraffin sections and immunocytochemistry. Acta Neuropathol. 2006;112(4):389–404. doi:10.1007/s00401-006-0127-z

38. Thal DR, Rüb U, Orantes M, Braak H. Phases of Aβ-deposition in the human brain and its relevance for the development of AD. Neurology. 2002;58(12):1791–1800. doi:10.1212/WNL.58.12.1791

39. Mirra SS, Heyman A, McKeel D, et al. The Consortium to Establish a Registry for Alzheimer’s Disease (CERAD): Part II. Standardization of the neuropathologic assessment of Alzheimer’s disease. Neurology. 1991;41(4):479–486. doi:10.1212/WNL.41.4.479

40. Fischer DL, Spina S, Miller BL, Seeley WW, Grinberg LT. FTLD-TDP versus LATE-NC: Experience of a Brain Bank specializing in FTLD-TDP. Alzheimers Dement Diagn Assess Dis Monit. 2026;18(3):e70411. doi:10.1002/dad2.70411

41. Nelson PT, Schmitt FA, Lin Y, et al. Hippocampal sclerosis in advanced age: clinical and pathological features. Brain. 2011;134(5):1506–1518. doi:10.1093/brain/awr053

42. McAleese KE, Alafuzoff I, Charidimou A, et al. Post-mortem assessment in vascular dementia: advances and aspirations. BMC Med. 2016;14(1):129. doi:10.1186/s12916-016-0676-5

43. Banerjee G, Carare R, Cordonnier C, et al. The increasing impact of cerebral amyloid angiopathy: essential new insights for clinical practice. J Neurol Neurosurg Psychiatry. 2017;88(11):982–994. doi:10.1136/jnnp-2016-314697

44. McKeith IG, Dickson DW, Lowe J, et al. Diagnosis and management of dementia with Lewy bodies: Third report of the DLB consortium. Neurology. 2005;65(12):1863–1872. doi:10.1212/01.wnl.0000187889.17253.b1

45. Crutch SJ, Schott JM, Rabinovici GD, et al. Consensus classification of posterior cortical atrophy. Alzheimers Dement. 2017;13(8):870–884. doi:10.1016/j.jalz.2017.01.014

46. Gorno-Tempini ML, Hillis AE, Weintraub S, et al. Classification of primary progressive aphasia and its variants. Neurology. 2011;76(11):1006–1014. doi:10.1212/WNL.0b013e31821103e6

47. Hammers DB, Eloyan A, Taurone A, et al. Profiling baseline performance on the Longitudinal Early-Onset Alzheimer’s Disease Study (LEADS) cohort near the midpoint of data collection. Alzheimers Dement. 2023;19(Suppl 9):S8–S18. doi:10.1002/alz.13160

48. Kramer JH, Jurik J, Sha SJ, et al. Distinctive Neuropsychological Patterns in Frontotemporal Dementia, Semantic Dementia, And Alzheimer Disease. Cogn Behav Neurol. 2003;16(4):211–218. doi:10.1097/00146965-200312000-00002

49. Ramos EM, Dokuru DR, Van Berlo V, et al. Genetic screen in a large series of patients with primary progressive aphasia. Alzheimers Dement. 2019;15(4):553–560. doi:10.1016/j.jalz.2018.10.009

50. Ramos EM, Dokuru DR, Van Berlo V, et al. Genetic screening of a large series of North American sporadic and familial frontotemporal dementia cases. Alzheimers Dement. 2020;16(1):118–130. doi:10.1002/alz.12011

51. Iaccarino L, La Joie R, Edwards L, et al. Spatial Relationships between Molecular Pathology and Neurodegeneration in the Alzheimer’s Disease Continuum. Cereb Cortex. 2021;31(1):1–14. doi:10.1093/cercor/bhaa184

52. Sled JG, Zijdenbos AP, Evans AC. A nonparametric method for automatic correction of intensity nonuniformity in MRI data. IEEE Trans Med Imaging. 1998;17(1):87–97. doi:10.1109/42.668698

53. Ashburner J, Friston KJ. Unified segmentation. NeuroImage. 2005;26(3):839–851. doi:10.1016/j.neuroimage.2005.02.018

54. Strom A, Iaccarino L, Edwards L, et al. Cortical hypometabolism reflects local atrophy and tau pathology in symptomatic Alzheimer’s disease. Brain. 2022;145(2):713–728. doi:10.1093/brain/awab294

55. Klunk WE, Koeppe RA, Price JC, et al. The Centiloid Project: Standardizing quantitative amyloid plaque estimation by PET. Alzheimers Dement. 2015;11(1):1–15.e1-4. doi:10.1016/j.jalz.2014.07.003

56. Lesman-Segev OH, La Joie R, Stephens ML, et al. Tau PET and multimodal brain imaging in patients at risk for chronic traumatic encephalopathy. NeuroImage Clin. 2019;24:102025. doi:10.1016/j.nicl.2019.102025

57. Royse SK, Lopresti BJ, Murphy A, et al. Validation of amyloid PET positivity thresholds in centiloids: a multisite PET study approach. Alzheimers Res Ther. 2021;13(1):99. doi:10.1186/s13195-021-00836-1

