## Supplementary Tables and figures for "Too early for LATE? Age at onset is associated with the clinical and neuroimaging expression of limbic-predominant age-related TDP-43 pathology in Alzheimer’s disease"

| **Supplementary Table 1 Clinical and neuropathologic characteristics according to LATE-NC status in early-onset cases** | | | |
|---|---|---|---|
| | **LATE-NC- (n = 100)** | **LATE-NC+ (n = 23)** | **P-value** |
| **Demographic and clinical characteristics** | | | |
| Age at symptom onset, years | 55.5 ± 5.8 | 58.5 ± 6.0 | 0.035 |
| Age at death, years | 66.2 ± 6.6 | 73.6 ± 6.4 | <0.001 |
| Duration from symptom onset to death, years | 10.5 ± 3.4 | 15.1 ± 4.7 | <0.001 |
| Female sex, n (%) | 48 (48%) | 16 (70%) | 0.073 |
| Education, years | 16.3 ± 3.5 | 18.1 ± 2.7 | 0.009 |
| Clinical phenotype, n (%) | | | 0.195 |
| Amnestic | 55 (59%) | 17 (81%) | |
| Non-amnestic* | 11 (12%) | 2 (9.5%) | |
| Primary progressive aphasia | 12 (13%) | 0 (0%) | |
| Posterior cortical atrophy | 16 (17%) | 2 (9.5%) | |
| Baseline MMSE | 20.8 ± 5.6 | 23.1 ± 5.9 | 0.105 |
| Baseline CDR Sum of Boxes | 4.8 ± 2.4 | 5.4 ± 2.8 | 0.37 |
| **Neuropathologic features** | | | |
| AD neuropathologic change (ADNC)**, n (%) | | | >0.999 |
| Intermediate | 3 (3%) | 1 (4%) | |
| High | 97 (97%) | 22 (96%) | |
| Lewy body disease, n (%) | 57 (57%) | 18 (78%) | 0.1 |
| Hippocampal sclerosis, n (%) | 3 (3%) | 4 (19%) | 0.02 |
| Cerebral amyloid angiopathy, n (%) | 80 (80%) | 20 (87%) | 0.56 |
| Vascular pathology, n (%) | 27 (27%) | 7 (30%) | 0.80 |

Data are presented as mean ± standard deviation for continuous variables and n (%) for categorical variables. Between-group comparisons were performed using Student's t-test for continuous variables and $\chi^2$ test for categorical variables. Percentages were calculated based on available data; denominators may vary because of missing data. *Non-amnestic phenotype included dysexecutive and other atypical presentations not classified as primary progressive aphasia or posterior cortical atrophy. ** One LATE-NC+ case had ADNC severity indeterminate between intermediate and high.

AD, Alzheimer's disease; LATE-NC, limbic-predominant age-related TDP-43 encephalopathy neuropathologic change.

| **Supplementary Table 2 APOE ε4 and TMEM106B genetic risk according to LATE-NC status in early-onset cases** | | | |
|---|---|---|---|
| | **LATE-NC- (n = 95)** | **LATE-NC+ (n = 21)** | **P-value** |
| *APOE* ε4 carrier, n (%) | 48/95 (51%) | 15/21 (71%) | 0.095 |
| *TMEM106B* rs1990622 risk genotype, n (%) | 25/93 (27%) | 10/21 (48%) | 0.072 |
| Both *APOE* ε4 carrier and *TMEM106B* rs1990622 A/A, n (%) | 16/93 (17%) | 8/21 (38%) | 0.043 |

Data are presented as n (%), and between-group comparisons were performed using Fisher's exact test. Percentages were calculated based on available data; denominators may vary because of missing data.
*APOE*, apolipoprotein; LATE-NC, limbic-predominant age-related TDP-43 encephalopathy neuropathologic change; *TMEM106B*, transmembrane protein 106B.

| **Supplementary Table 3 Distribution of LATE-NC stage by age at symptom onset and age at death** | | | | |
|---|---|---|---|---|
| **Age group, years** | **Stage 1, n (%)** | **Stage 2, n (%)** | **Stage 3, n (%)** | **Total** |
| **Age at symptom onset** | | | | |
| ≤55 | 1 (12.5%) | 7 (87.5%) | 0 | 8 |
| 56–65 | 5 (33.3%) | 10 (66.7%) | 0 | 15 |
| 66–75 | 3 (13.0%) | 19 (82.6%) | 1 (4.3%) | 23 |
| ≥76 | 2 (14.3%) | 12 (85.7%) | 0 | 14 |
| **Age at death** | | | | |
| ≤65 | 0 | 1 (100%) | 0 | 1 |
| 66–75 | 3 (23.1%) | 10 (76.9%) | 0 | 13 |
| 76–85 | 5 (18.5%) | 22 (81.5%) | 0 | 27 |
| ≥86 | 4 (20.0%) | 15 (75.0%) | 1 (5.0%) | 20 |

Data are presented as n (%), with percentages calculated within each age group. Age at symptom onset was unavailable for one individual with LATE-NC.
LATE-NC, limbic-predominant age-related TDP-43 encephalopathy neuropathologic change

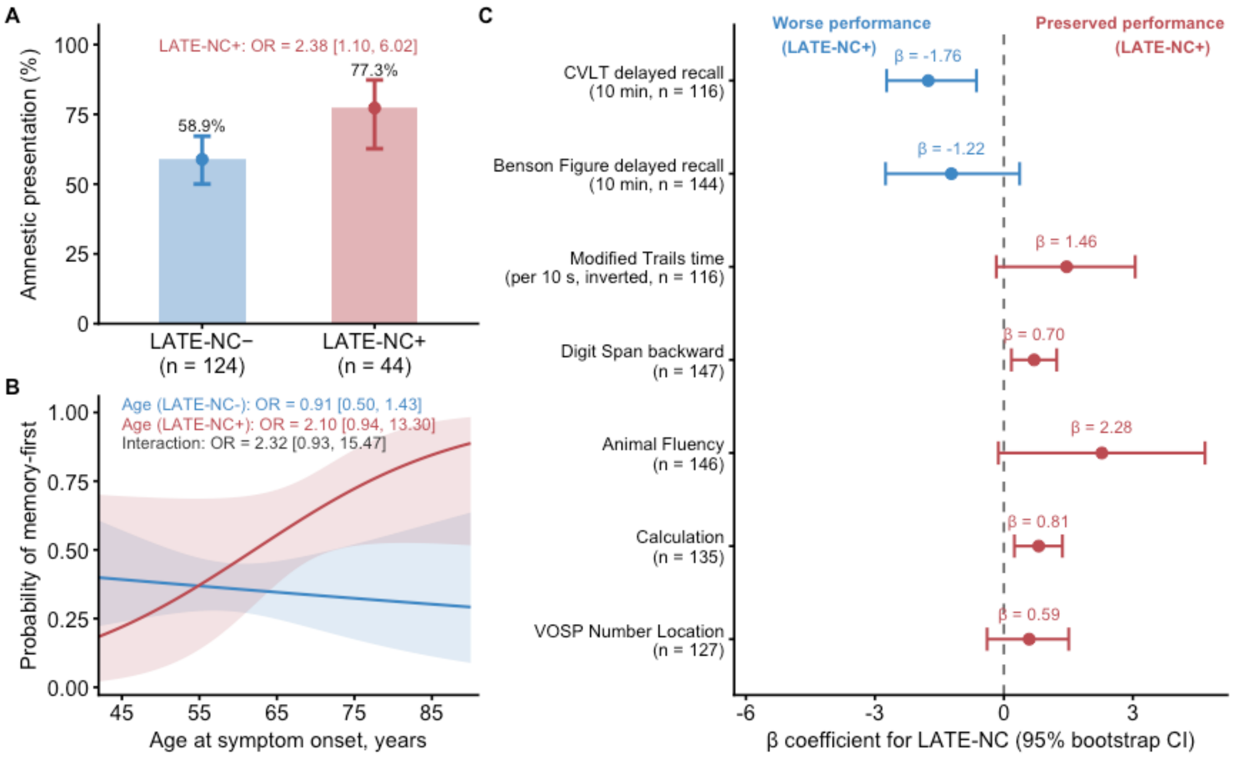


**Supplementary Figure 1 Sensitivity analysis of clinical presentation and cognitive performance according to LATE-NC status after exclusion of LATE-NC stage 1.** (A) Observed proportion of individuals with an amnestic presentation at first evaluation according to LATE-NC status. Bars represent observed proportions, points and error bars represent model-estimated probabilities and their 95% CIs. (B) Model-estimated probability of memory being the first reported symptom across age at symptom onset, stratified by LATE-NC status. Shaded areas represent 95% CIs. (C) Associations between LATE-NC and cognitive performance at first evaluation. Points represent β coefficients for LATE-NC and error bars represent 95% CIs. Modified Trails time was inverted so that higher values indicate better performance. Models were adjusted for age at symptom onset, with sex and education retained when they improved model fit, 95% CIs were estimated using 5,000 bootstrap resamples. The Boston Naming Test was also evaluated, but LATE-NC was not retained in the final AIC-selected model and therefore is not shown.

CI, confidence interval; CVLT, California Verbal Learning Test; LATE-NC, limbic-predominant age-related TDP-43 encephalopathy neuropathologic change; VOSP, Visual Object and Space Perception.

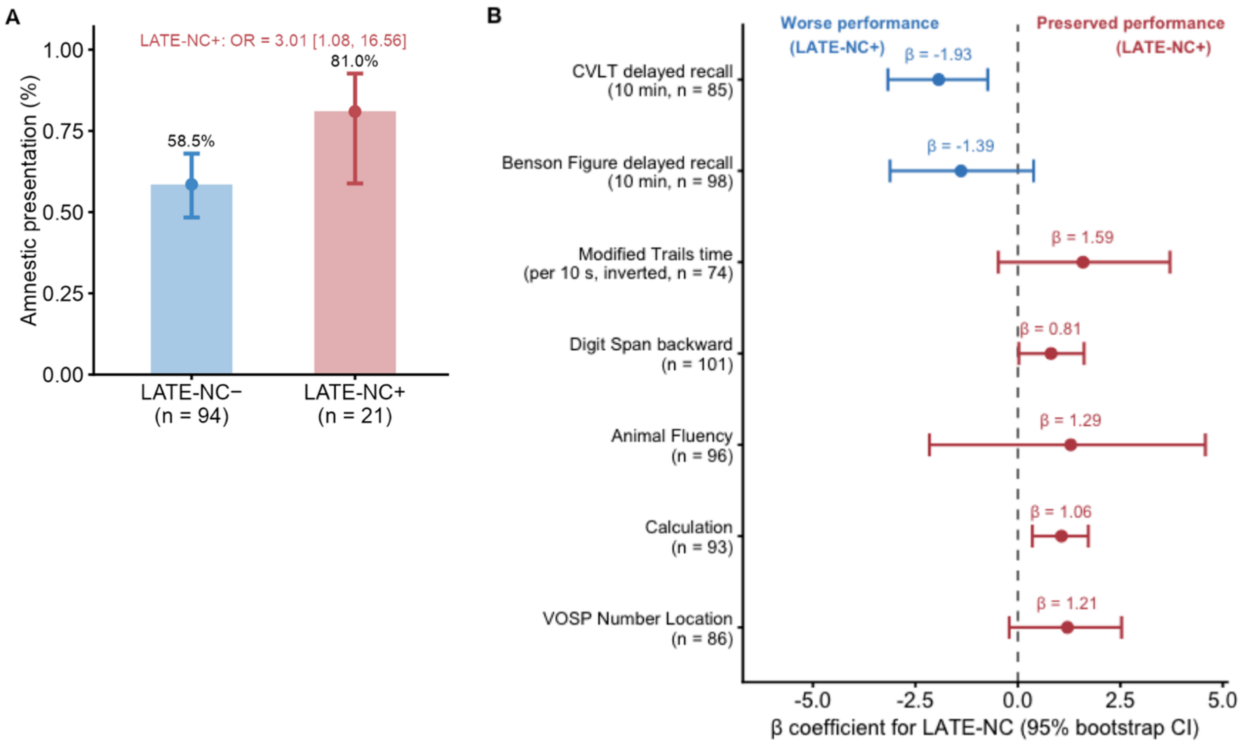


**Supplementary Figure 2 Sensitivity analysis of clinical presentation and cognitive performance according to LATE-NC status, restricted to early-onset cases.** (A) Observed proportion of individuals with an amnestic presentation at first evaluation according to LATE-NC status. Bars represent observed proportions, points and error bars represent model-estimated probabilities and their 95% CIs (B) Associations between LATE-NC and cognitive performance at first evaluation. Points represent β coefficients for LATE-NC and error bars represent 95% CIs. Modified Trails time was inverted so that higher values indicate better performance. Models were adjusted for age at symptom onset, with sex and education retained when they improved model fit, 95% CIs were estimated using 5,000 bootstrap resamples. The Boston Naming Test was also evaluated, but LATE-NC was not retained in the final AIC-selected model and therefore is not shown.

CI, confidence interval; CVLT, California Verbal Learning Test; LATE-NC, limbic-predominant age-related TDP-43 encephalopathy neuropathologic change; VOSP, Visual Object and Space Perception.

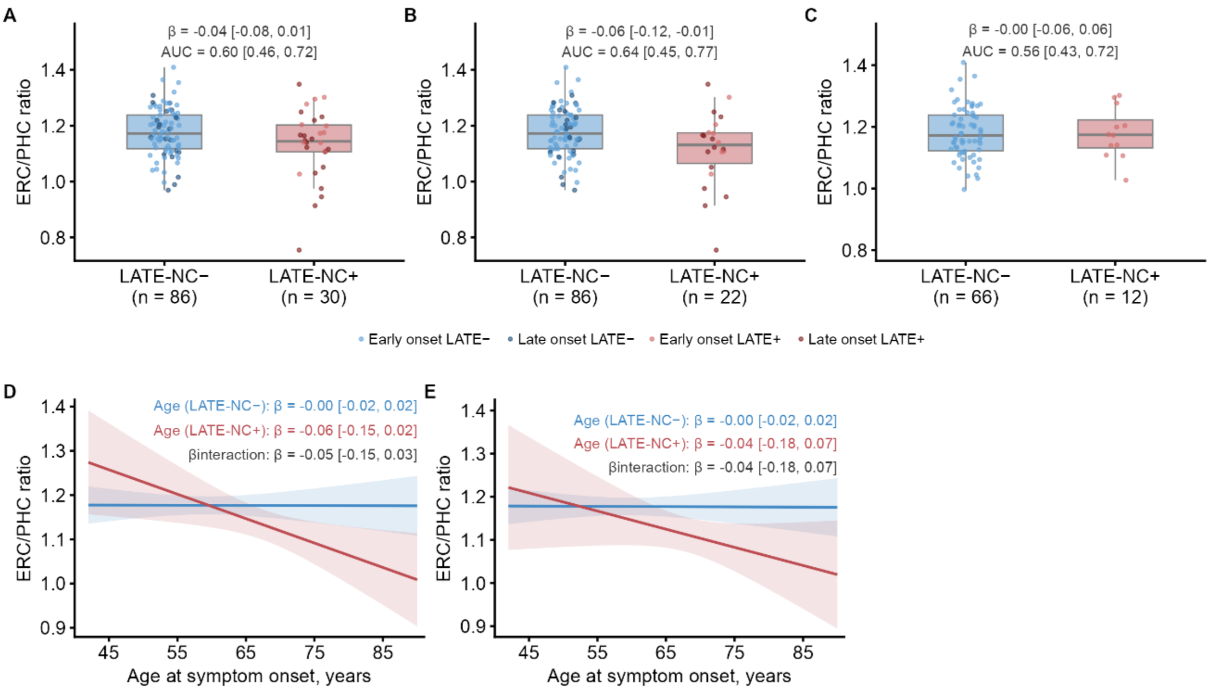


**Supplementary Figure 3 Entorhinal-to-parahippocampal (ERC/PHC) volume ratio according to LATE-NC status and age at symptom onset.** (A) ERC/PHC volume ratio by LATE-NC status in the primary analysis, (B) after exclusion of LATE-NC stage 1, and (C) restricted to early-onset cases. Boxplots show the median and interquartile range, with individual observations in A-B stratified by early- and late-onset cases. β coefficients (95% CI) reflect the association between LATE-NC and ERC/PHC ratio; AUC values reflect the discriminative performance of the ERC/PHC ratio for LATE-NC status. (D) Model-estimated ERC/PHC ratio across age at symptom onset by LATE-NC status in the primary analysis and (E) after exclusion of LATE-NC stage 1. Lines represent model estimates and shaded areas represent 95% CIs. Models were adjusted for age at symptom onset and total intracranial volume, with 95% CIs estimated using 5,000 bootstrap resamples.

AUC, area under the curve; CI, confidence interval; ERC, entorhinal cortex; LATE-NC, limbic-predominant age-related TDP-43 encephalopathy neuropathologic change; PHC, parahippocampal cortex.

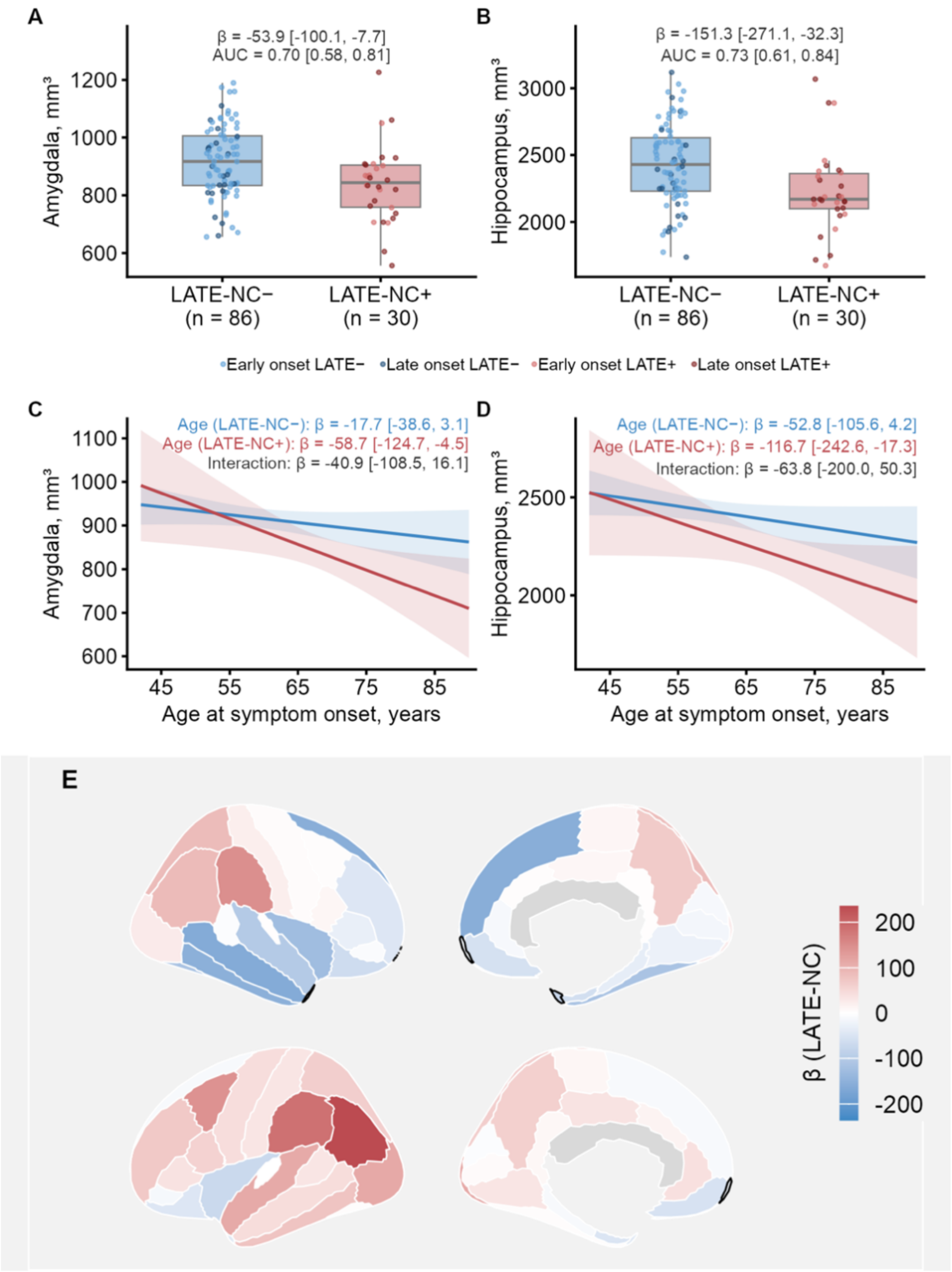


**Supplementary Figure 4 Sensitivity analysis of regional gray matter volumes according to LATE-NC status and age at symptom onset after exclusion of LATE-NC stage 1.** (A) Amygdala and (B) hippocampal gray matter volumes by LATE-NC status. Boxplots show the median and interquartile range, with individual observations stratified by early- and late-onset cases. β coefficients (95% CI) correspond to the association between LATE-NC and regional volumes; AUC values reflect the discriminative performance of amygdala and hippocampal volumes for LATE-NC status. (C) Amygdala and (D) hippocampal model-estimated volumes across age at symptom onset, by LATE-NC status. Lines represent model estimates and shaded areas represent 95% CIs. (E) Cortical gray matter volume differences associated with LATE-NC (LATE-NC+ vs. LATE-NC-). Upper and lower views correspond to the right and left hemispheres, respectively. Dark outlines mark regions where the 95% CIs did not include zero. Models were adjusted for age at symptom onset and total intracranial volume, with 95% CIs estimated using 5,000 bootstrap resamples.

AUC, area under the curve; CI, confidence interval; LATE-NC, limbic-predominant age-related TDP-43 encephalopathy neuropathologic change.

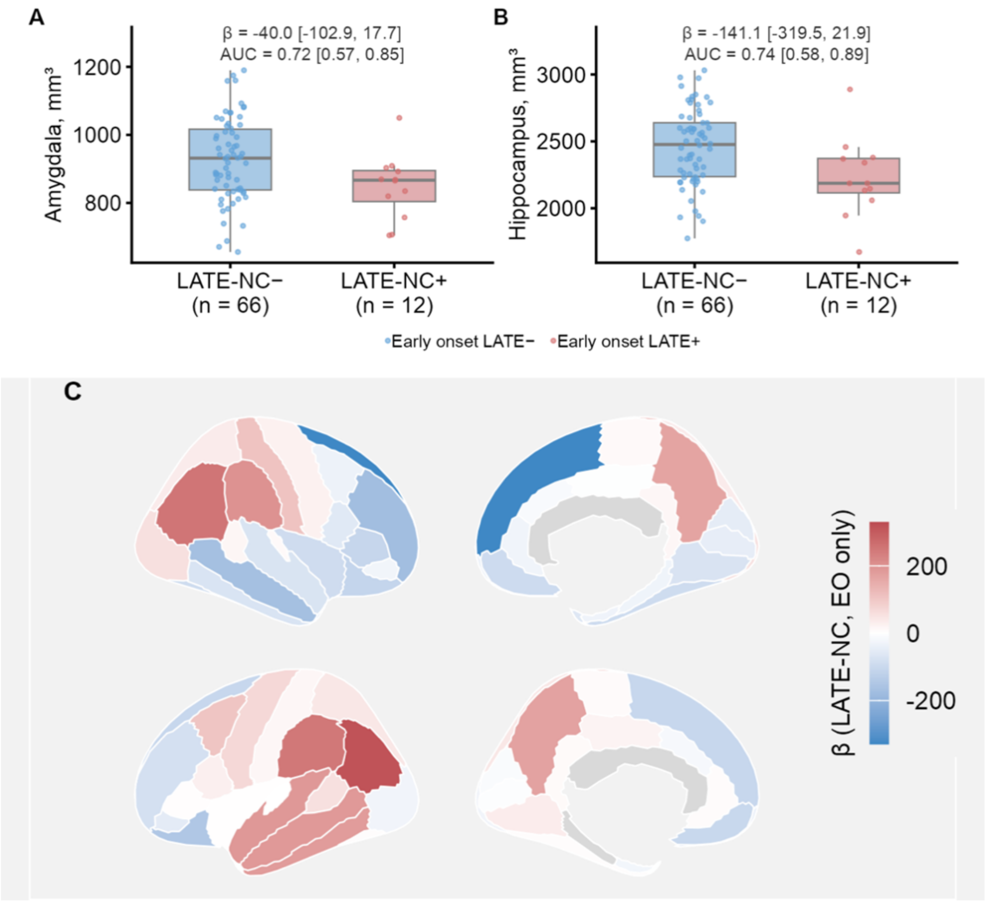


**Supplementary Figure 5 Sensitivity analysis of regional gray matter volumes according to LATE-NC status, restricted to early-onset cases.** (A) Amygdala and (B) hippocampal gray matter volumes by LATE-NC status. Boxplots show the median and interquartile range, with individual observations stratified by early- and late-onset cases. β coefficients (95% CI) correspond to the association between LATE-NC and regional volumes; AUC values reflect the discriminative performance of amygdala and hippocampal volumes for LATE-NC status. (C) Cortical gray matter volume differences associated with LATE-NC (LATE-NC+ vs. LATE-NC-). Upper and lower views correspond to the right and left hemispheres, respectively. Models were adjusted for age at symptom onset and total intracranial volume, with 95% CIs estimated using 5,000 bootstrap resamples.

AUC, area under the curve; CI, confidence interval; LATE-NC, limbic-predominant age-related TDP-43 encephalopathy neuropathologic change.

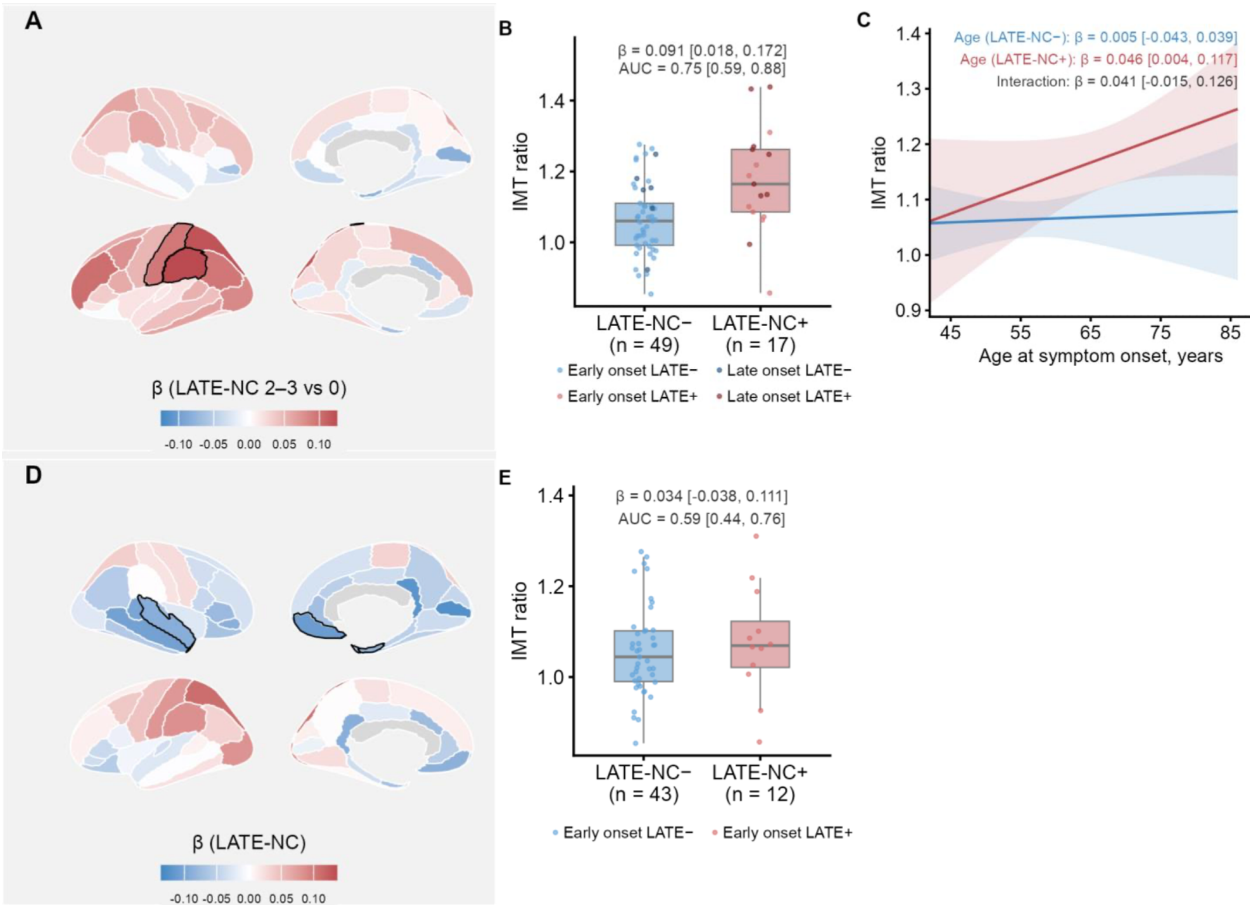


**Supplementary Figure 6 Sensitivity analyses of regional FDG-PET metabolism according to LATE-NC status and age at symptom onset.** (A-C) After exclusion of LATE-NC stage 1. (A) Cortical metabolic differences associated with LATE-NC (LATE-NC+ vs. LATE-NC-). Upper and lower views correspond to the right and left hemispheres, respectively. Dark outlines mark regions where the 95% CIs did not include zero. (B) Inferior-to-medial temporal (IMT) ratio by LATE-NC status. Boxplot shows the median and interquartile range, with individual observations stratified by early- and late-onset cases. β coefficients (95% CI) correspond to the association between LATE-NC and IMT ratio; AUC value reflects the discriminative performance of the IMT ratio for LATE-NC status. (C) Model-estimated IMT ratio across age at symptom onset by LATE-NC status. Lines represent model estimates and shaded areas represent 95% CIs. (D-E) Restricted to early-onset cases. (D) Cortical metabolic differences associated with LATE-NC in early-onset subgroup. Views and dark outlines are as described for (A). (E) IMT ratio by LATE-NC status in the early-onset group. Boxplot, β coefficients, and AUC are as described for (B).

AUC, area under the curve; CI, confidence interval; FDG-PET, $^{18}$F-fluorodeoxyglucose positron emission tomography; IMT, inferior-to-medial temporal; LATE-NC, limbic-predominant age-related TDP-43 encephalopathy neuropathologic change.

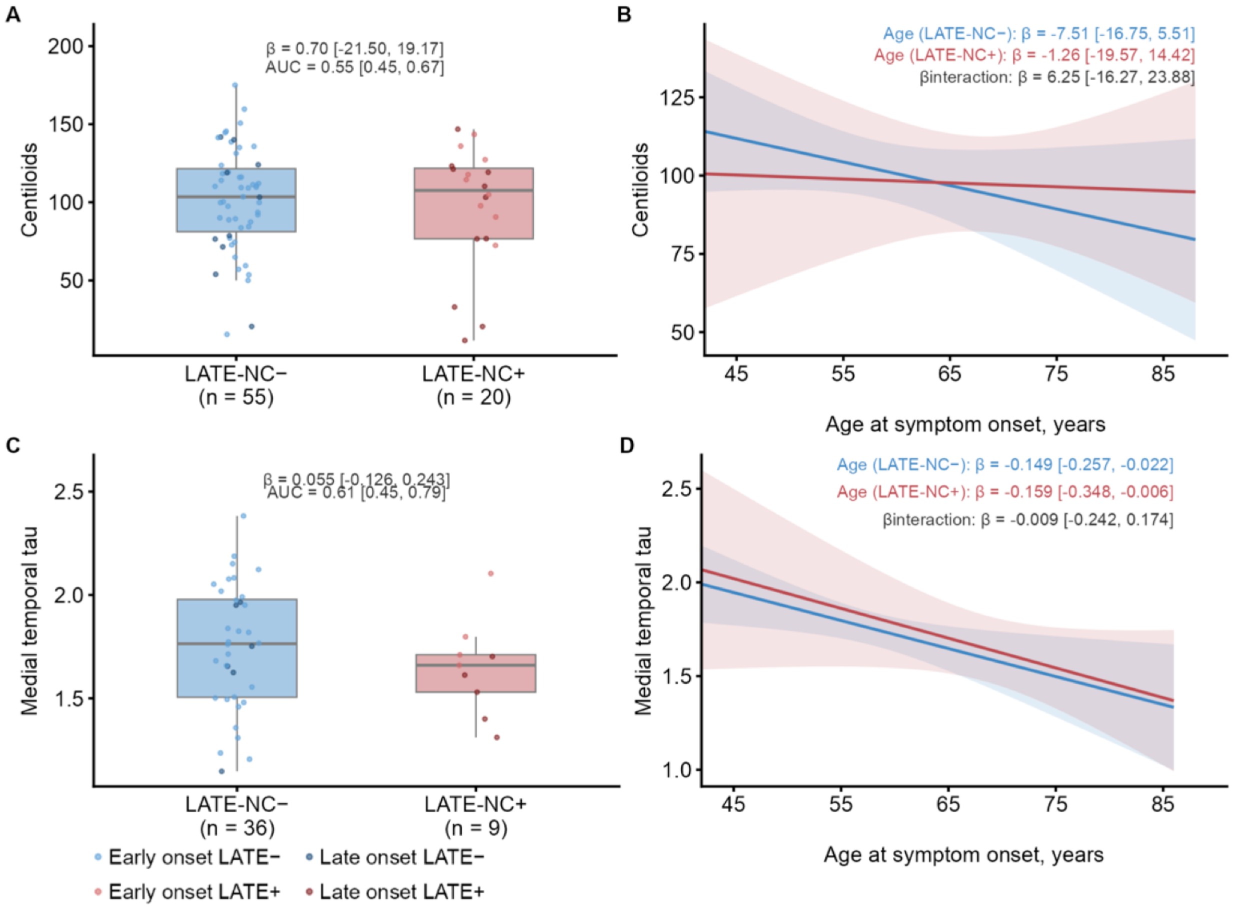


**Supplementary Figure 7 Amyloid and medial temporal tau-PET according to LATE-NC status and age at symptom onset**. (A) Amyloid burden, measured in Centiloids, by LATE-NC status. Boxplots show the median and interquartile range, with individual observations stratified by early- and late-onset cases. β coefficients (95% CI) correspond to the association between LATE-NC and Centiloid values for LATE-NC status; AUC value reflects the discriminative performance of the Centiloids for LATE-NC status. (B) Model-estimated Centiloid values across age at symptom onset by LATE-NC status. Lines represent model estimates and shaded areas represent 95% CIs. (C-D) Medial temporal tau-PET uptake, as described for (A-B). Models were adjusted for age at symptom onset, with 95% CIs estimated using 5000 bootstrap resamples.

CI, confidence interval; LATE-NC, limbic-predominant age-related TDP-43 encephalopathy neuropathologic change.
